# Combined family history and polygenic score prediction of major depressive disorder

**DOI:** 10.1101/2025.09.22.25336356

**Authors:** Rujia Wang, Helena L. Davies, Sang-Hyuck Lee, Johan Zvrskovec, Christopher Hübel, Saakshi Kakar, Chelsea Malouf, Laura Meldrum, Yuhao Lin, Iona Smith, Gursharan Kalsi, Henry C. Rogers, Shannon Bristow, Molly R. Davies, Donald M. Lyall, Allan H. Young, Anthony J. Cleare, Katrina A. S. Davis, Roland Zahn, Victor A. Gault, Le Roy C. Dowey, Ruth K. Price, Keith G. Thomas, James T. R. Walters, Daniel J. Smith, Chérie Armour, Ian R. Jones, Nathalie Kingston, John R. Bradley, Andrew M. McIntosh, NIHR BioResource consortium, The GLAD Study, Matthew Hotopf, Jonathan R.I. Coleman, Evangelos Vassos, Thalia C. Eley, Raquel Iniesta, Gerome Breen

## Abstract

**Importance:** Major depressive disorder (MDD) is a complex psychiatric disorder influenced by genetic, social, and environmental factors. Family history, genome-wide polygenic scores, and childhood trauma are key predictors of MDD onset with distinct contributions.

**Objective:** This study modelled the combined effects of family history of multiple psychiatric disorders, polygenic scores of multiple traits, and childhood trauma, alongside sociodemographic factors on MDD diagnosis and number of episodes. We aimed to build and externally validate predictive models for MDD risk and severity.

**Participants:** We used data from the Genetic Links to Anxiety and Depression Study and other NIHR BioResource studies (GLAD+) and UK Biobank (UKB) collected between 2016 and 2023.

**Outcomes and measurements:** MDD diagnosis followed DSM-5 criteria using online questionnaire data. Family history (Yes/No) was reported for up to 22 psychiatric disorders. Polygenic scores were calculated based on genome-wide association studies (n=22). Participants answered the five-item childhood trauma screener. We used elastic net regression with nested cross-validation to select the best predictors.

**Results:** In GLAD+ (9,927 MDD cases, 4,452 controls), family history explained 17% of the MDD variance, followed by childhood trauma (11%), sociodemographics (10%), and polygenic scores (7%), resulting in 33% of the MDD variance (AUC-ROC=0.84). In UKB (40,667 MDD cases, 70,755 controls), family history explained 13% of the MDD variance, childhood trauma (7%), sociodemographics (6%), and polygenic scores (4%). Combined together, the predictors explained 23% of the variance (AUC-ROC=0.74). The top five individual predictors were family history of depression, childhood trauma, female sex, family history of anxiety, and the MDD polygenic score in both cohorts. The predictive models were externally well-validated across GLAD+ and UKB obtaining comparable predictive performance. Finally, when combined, the predictors explained 26% and 12% of the variance in the number of MDD episodes in GLAD+ and UKB respectively.

**Conclusions:** Integrating family history, PRSs, ChT, and sociodemographic factors can predict risk of an MDD diagnosis and its recurrent course. This model may help identify individuals at high risk for depression in clinical settings, assess its severity, enable early diagnosis and potentially personalize treatment.

## Introduction

Major depressive disorder (MDD) is one of the most common psychiatric disorders worldwide, with a lifetime prevalence of 15-18%^1^. Around 35-75% of individuals with an initial episode may have recurrent episodes^2–6^. MDD reduces social functioning and work ability, diminishes health-related quality of life, and elevates suicide risk^7–9^. Globally, depression ranked second among the top 25 leading causes of years lived with disability and contributed 45.7 million disability-adjusted life-years in 2019^10,11^. Genetic, social, environmental, and sociodemographic risk factors contribute to the development of depression^12^. Combining these risk factors may increase accuracy of prediction of MDD risk and recurrence, and thus early diagnosis.

The past decade has seen increasingly successful genome-wide association studies (GWASs) of depression^13–16^. A 2025 genome-wide meta-analysis identified >600 independent depression-associated loci^16^. Furthermore, polygenic risk scores (PRSs) for depression now explain 1.5-3.2% of its variance^14^. However, integrating genomics into clinical prediction remains one of the top challenges in psychiatry^17^. Although PRS captures more precise genetic effects than family history, it only accounts for the common genetic variants identified in a GWAS, making it insufficient by itself for prediction. In contrast, family history of depression is a strong and robust risk factor both for the development of depression^18–21^, and the severity and recurrence of illness^22–24^. The offspring of those diagnosed with MDD are two-to-three times more likely to develop MDD^25^. Many have advocated for the wider use of family history in primary care for prevention^26,27^. Childhood trauma (ChT) is another major risk factor for depression^28–30^. A meta-analysis of 184 studies found that those who experienced childhood trauma were 2.7-3.7-times more likely to develop depression in adulthood, with an earlier onset and twice the likelihood of developing chronic or treatment-resistant depression^29^. Additional established predictors include sex and age^31^: females are about twice as likely as males to develop MDD^32^ and the prevalence of MDD increases during adolescence and young adulthood, remains stable in adulthood, and declines in middle age^33^.

Studies in psychiatry and other fields^34–37^ apply supervised machine learning^38,39^ combining multiple PRSs for outcome prediction^38,39^, leveraging genetic overlap across psychiatric disorders and other traits to maximise predictive power^40^. Supervised machine learning is used to predict outcomes like risk and prognosis of MDD incorporating predictors. Machine learning relies on patterns within the data, enabling a bottom-up, data-driven approach^37^, making predictions without presupposing causal relationships between predictors and outcomes^41^. The primary objective is to maximise predictive accuracy, irrespective of the size and direction of potential causal associations^42,43^.

PRSs alone have limited predictive power. Therefore, we used machine learning to test the extent to which family history of multiple psychiatric disorders (mFH), PRSs of multiple psychiatric disorders and related traits (mPRS), childhood trauma, and sociodemographic factors could predict the risk, recurrence and severity of MDD. Models were built with data from the Genetic Links to Anxiety and Depression Study and other NIHR BioResource cohorts (GLAD+) and externally validated in data from UK Biobank (UKB) and vice-versa.

## Methods

### Study sample

Participants were recruited into the GLAD+ Study, which combines two UK cohorts: the GLAD Study (www.gladstudy.org.uk)^44^, and the NIHR BioResource COVID-19 Psychiatry and Neurological Genetics (COPING) Study. The GLAD Study recruits participants with depression and/or anxiety. Participants provide demographic, environmental, and genetic data and consent to medical record linkage and recontact. It includes >64,000 consented participants, with >50,000 having completed online surveys, and >35,000 saliva samples. During the COVID-19 pandemic, the GLAD team recontacted GLAD participants and healthy volunteers from other NIHR BioResource (https://bioresource.nihr.ac.uk/) studies to conduct the COPING study, including >20,000 participants with psychiatric disorders and >11,000 healthy volunteers, two-thirds of whom have been genotyped. Most MDD cases (78%) in the current study were originally enrolled in the GLAD Study^44^, while all controls were from the COPING study (details, see Supplement). The GLAD Study was approved by the London - Fulham Research Ethics Committee on 21st August 2018 (REC reference: 18/LO/1218) following a full review by the committee. The NIHR BioResource has been approved as a Research Tissue Bank by the East of England - Cambridge Central Committee (REC reference: 17/EE/0025).

The UKB is a prospective health study of >500,000 UK individuals, aged 40-69 at recruitment (2006-2010). In 2016-17, participants were invited via email and postal newsletter to complete the first online Mental Health Questionnaire (MHQ1), with >157,000 responses by July 2017^45^. In 2022, the second Mental Health Questionnaire (MHQ2) was distributed, leading to >169,000 responses. Over 206,000 UK Biobank participants have data from either MHQ1 or MHQ2. In MHQ2, participants reported family history of psychiatric disorders in first-degree relatives. All participants provided informed consent.

### Measurements

#### Major depressive disorder (MDD)

Lifetime major depressive disorder was assessed using the Adapted Composite International Diagnostic Interview-Short Form (CIDI-SF), based on DSM-5 criteria in both studies^46^. The GLAD+ study included 20,191 MDD cases and 11,064 controls. The UKB included 40,667 MDD cases and 70,755 controls (definitions in **Table S1**).

Both studies assessed the number of depression periods lasting two or more weeks, with 13 answer options ranging from “1” to “13 or more”. Age of onset was determined by asking the participant’s age at their first two-week depression period. Treatment-resistant depression in GLAD+ was assessed using the 4-item Maudsley Staging Method^47^ (scored 0-10, **Table S2**) and the Patient Health Questionnaire-9^48^ current depression severity scale (recoded 0-4), with the degree of treatment-resistant scores ranging from 0 to 14.

#### Family history (FH)

In GLAD+, family history of psychiatric disorders was assessed by asking if any family members had ever been diagnosed with each of 22 psychiatric disorders (**Table S3**, optional survey). In UKB, family history was assessed by asking if any first-degree blood relatives had ever been diagnosed with each of nine psychiatric disorders (**Table S4**, MHQ2). Participants reporting these conditions were considered to have a family history of the corresponding mental health disorders.

#### Childhood trauma (ChT)

Childhood trauma was assessed using the five-item Likert-scale Childhood Trauma Screen^49^, covering emotional, physical, and sexual abuse, and emotional and physical neglect. Participants reporting any type of trauma were classified as having experienced childhood trauma (**Table S5**).

#### MegaPRS

We generated PRSs using GWAS summary statistics excluding UK or UKB samples for MDD, anxiety, ADHD, bipolar disorder, ASD, PTSD, and BMI (**Table S6**). We also generated PRSs for 15 additional psychiatric traits in the GLAD+ study, using summary statistics including UKB samples (**Table S6**). We used the GenoPred pipeline^50^ calculating MegaPRS^51^ based on the LDAK heritability model, where the variance explained by each SNP depends on its allele frequency, linkage disequilibrium, and functional annotations.

### Statistical analysis

We used univariable logistic regression to estimate MDD variance explained. We built a predictive model using multivariable nested elastic net regression^52^, which prevents overfitting and manages multicollinearity^40^. Nested regularisation improved model selection and generalization. We performed repeated nested cross-validation by splitting data into four folds for training, with ten-fold inner cross-validation to tune parameters, and one fold for testing. This process was repeated across all folds (**Figure 1**). Test predictions were compared to true outcomes, and accuracy for classification was assessed using ROC-AUC (receiver operating characteristic curve area under curve) or the lowest mean squared error^53^. Class weighting adjusted for case-control imbalance (details in Supplement).

**Figure 1.**
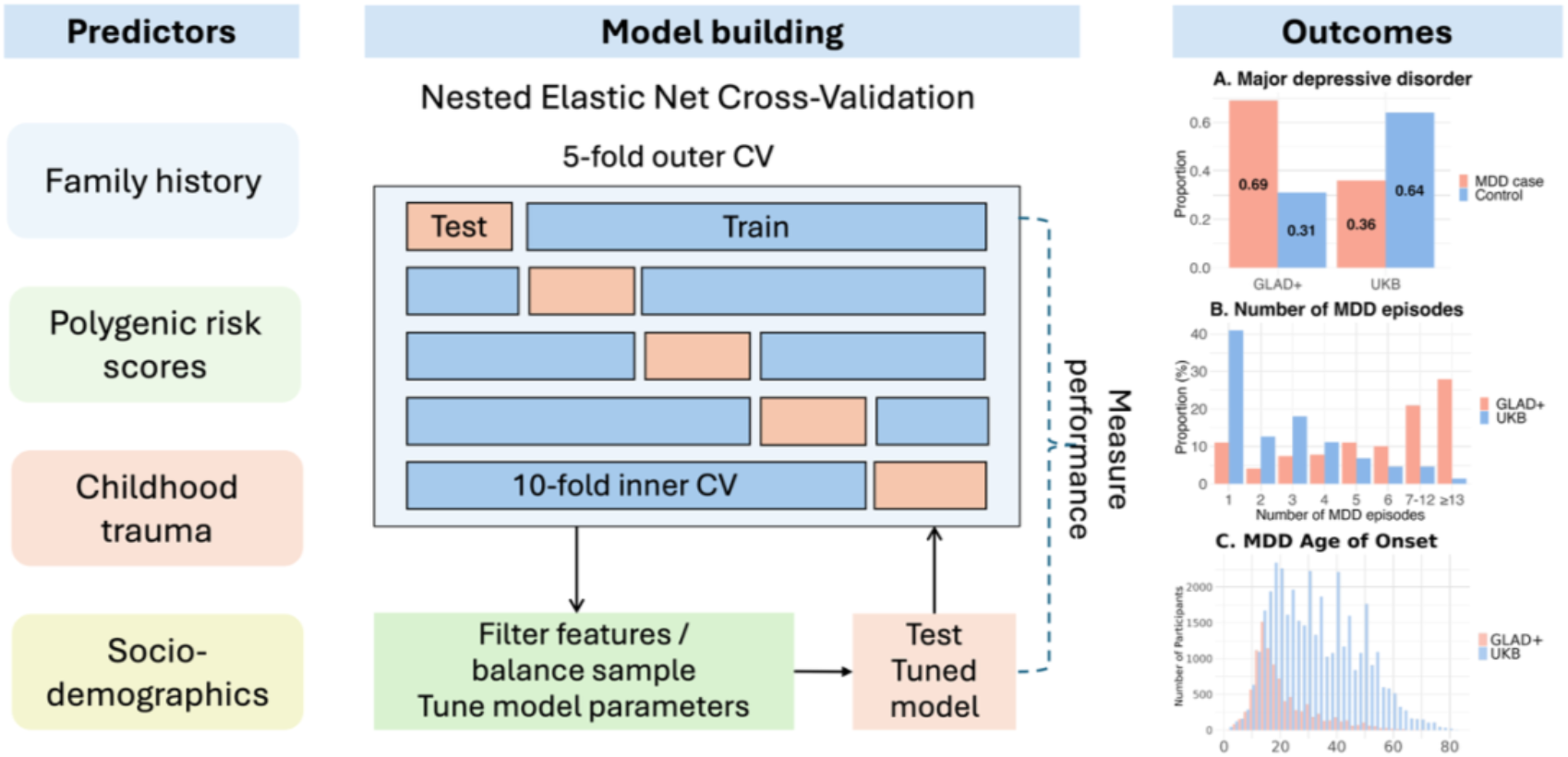
Flow chart of prediction model. Predictors include family history of multiple psychiatric disorders, polygenic risk scores for multiple psychiatric disorders and related traits, childhood trauma, and sociodemographics (sex and 10 genetic principal components). We performed repeated nested cross-validation, splitting the data into four folds for training, with ten-fold inner cross-validation to tune model parameters, and one fold for testing. This process was repeated across all folds. Outcomes include MDD diagnosis, number of episodes, and age of onset in GLAD+ and UKB. The model was trained on one dataset and externally validated on another, and vice versa.

The GLAD+ model included four predictor groups: (1) Family history of 22 psychiatric disorders (mFH), (2) PRSs of 11 psychiatric disorders and 11 related traits (mPRS), (3) Childhood trauma (ChT), and (4) Sociodemographic factors (sex assigned at birth and population stratification, modeled by ten genetic principal components [PCs]). Predictors were mapped across GLAD+ and UKB (**Table S4**), including mFH for nine psychiatric disorders, mPRS for seven disorders and traits, childhood trauma, and sociodemographic factors (sex and ten PCs).

We trained three models in GLAD+: (1) MDD diagnosis, (2) number of MDD episodes, and (3) age of onset, then tested them in UKB, and vice versa. MDD episode distributions differed between GLAD+ (severe cases) and UKB (mild cases), so we resampled the UKB MDD cases to match the GLAD+ distribution (n=2,442). We also tested the model built for MDD diagnosis to predict three MDD features: number of episodes, age of onset, and the degree of treatment-resistance. The first two were tested in both datasets, whilst treatment-resistance was only tested in GLAD+.

The model’s overall variance and each predictor group’s (i.e., mFH, mPRS, ChT, sociodemographics) contribution were quantified using the coefficient of determination (R^2^), with nested regularization ensuring balanced variance across groups. Liability R^2^ was converted using a severe MDD prevalence of 5% in GLAD+ and a mild MDD prevalence of 15% in UKB. We calculated positive predictive value (PPV), negative predictive value (NPV), sensitivity, and specificity at various prediction score cut-offs (0.3-0.7) to assess MDD classification. Calibration plots evaluated model fitness by comparing predicted probabilities with actual outcomes. All analyses were performed using R v4.3.0^54^.

## Results

Compared with controls (GLAD+ = 4,452; UKB = 70,755), participants with MDD (GLAD+ = 9,927; UKB = 40,667) were generally younger, female, more likely to have a family history of depression and anxiety, and experienced more childhood trauma (**Table S4**). The GLAD+ study had more severe MDD cases, with 89% experiencing recurrent episodes and 28% reporting 13 or more episodes. In contrast, 41% of UKB MDD cases had a single episode, and 72% had 3 episodes or less (**Figure 2**).

**Figure 2.**
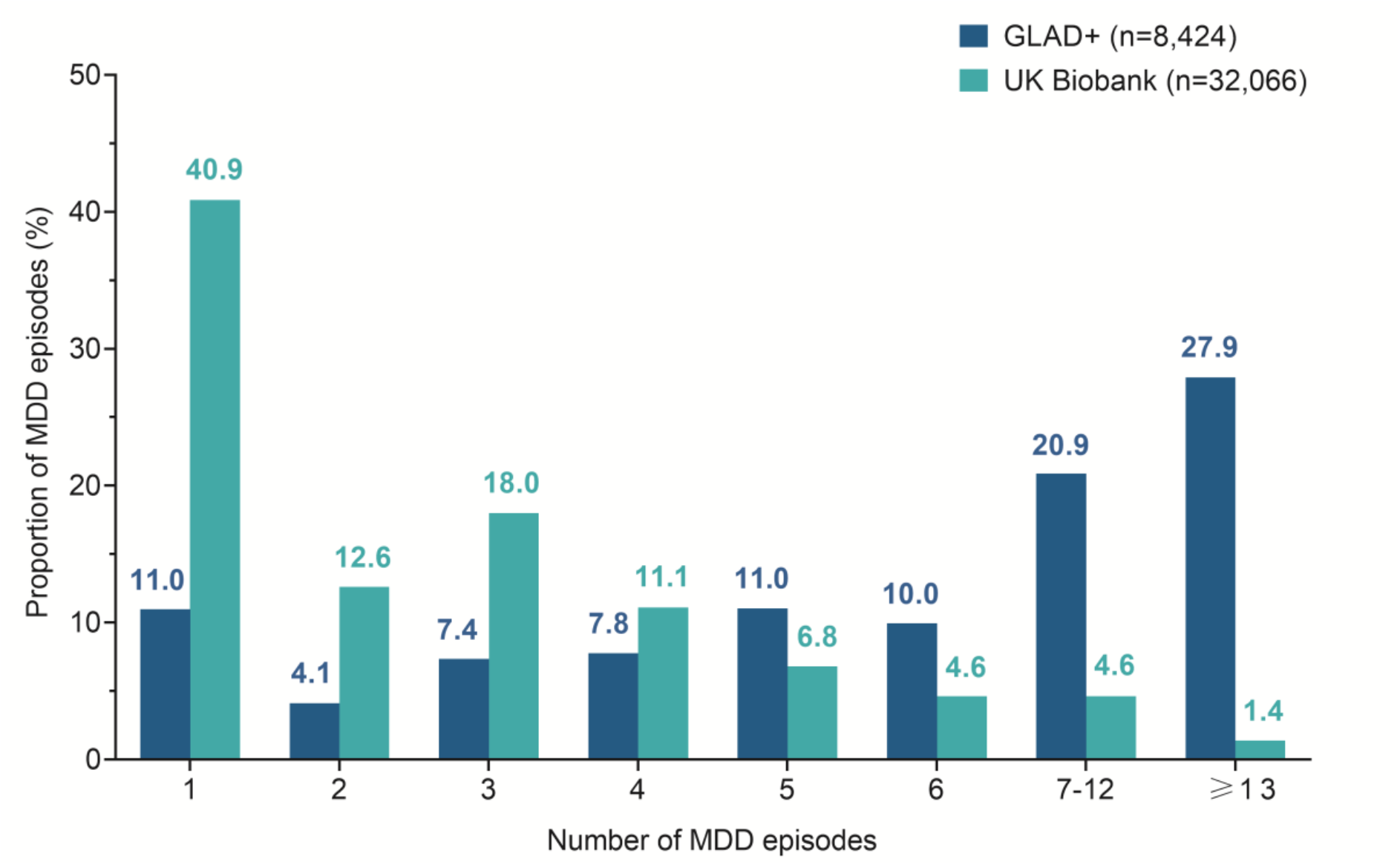
The number of MDD episodes in GLAD+ and UK.

Total variance explained was 33% in GLAD+ and 23% in UKB, by predictor group: mFH (17% vs 13%), ChT (11% vs 7%), demographics (10% vs 6%), and mPRS (7% vs 4%) (**Figure 3**). **Figure S1A-B** shows that mPRS added 2-5% points of variance beyond mFH alone. Additionally, compared to the best individual predictive utility of a family history of depression (14%) and a PRS of MDD (6%), both mFH and mPRS accounted for a greater quantity of MDD variance (**Figure S2**).

**Figure 3.**
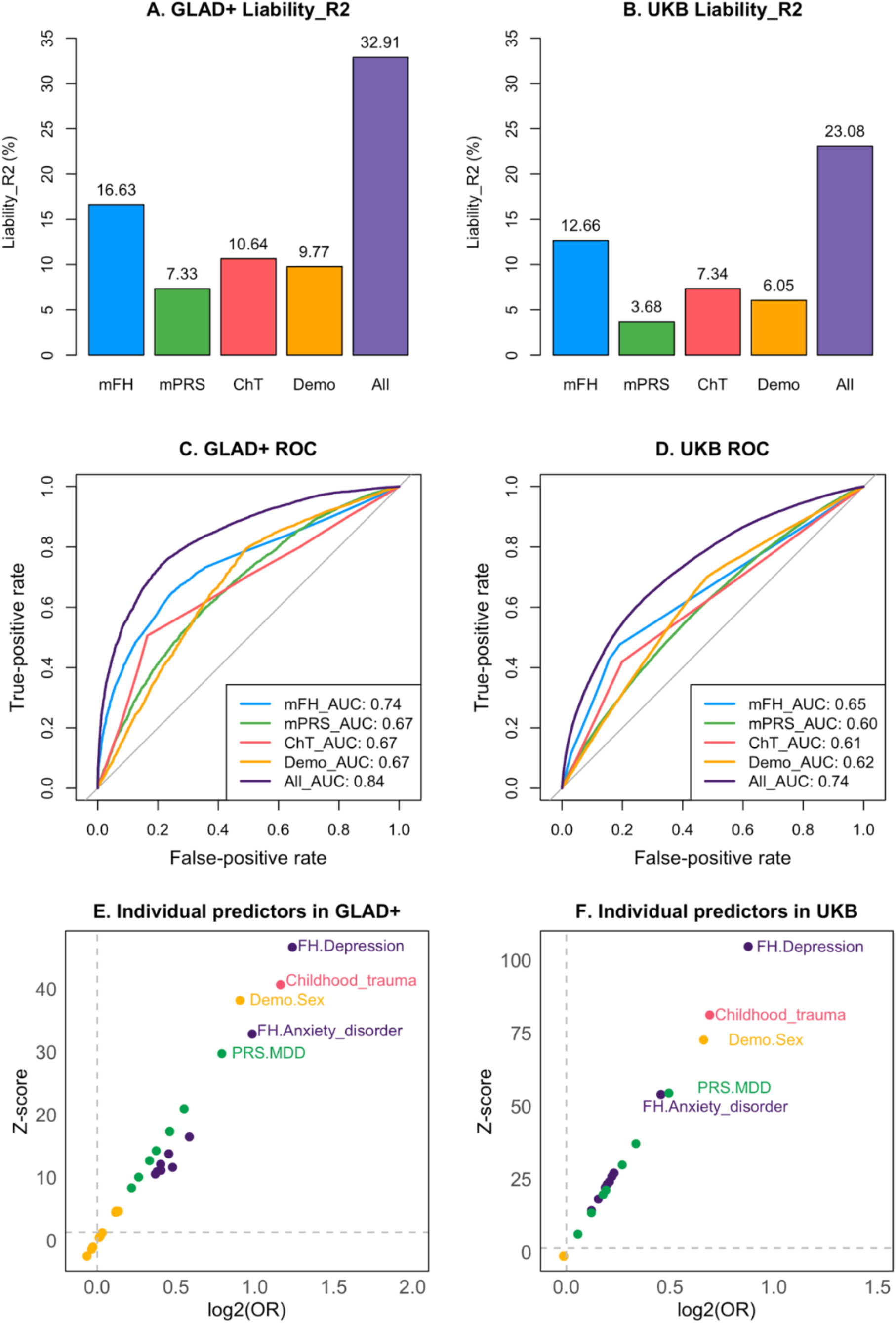
The effect of group and individual predictors for MDD in GLAD+ and UKB. The liability R^2^ of MDD explained by each group of predictors in GLAD+ (**Figure 3A**), with a severe MDD prevalence of 5%, and in UKB (**Figure 3B**), with a mild MDD prevalence of 15%. The ROC of each group of predictors in GLAD+ (**Figure 3C**) and in UKB (**Figure 3D**). mFH, family history of multiple psychiatric disorders, including family history of 22 psychiatric disorders in GLAD+, and family history of 9 psychiatric disorders in UKB. mPRS, polygenic risk scores of 11 psychiatric disorders and 11 related traits in GLAD+, and seven PRSs of psychiatric disorders and BMI based without UK/UKB GWAS sumstats in UKB. ChT, childhood trauma. Demo, sociodemographic factors, including sex and 10 genetic principal components (modelling population stratification). All, including all groups of predictors. ROC, receiver operating characteristic curve. Individual predictor effects for MDD in GLAD+ (**Figure 3E**) and UKB (**Figure 3F**), with all individual predictors mapped across both datasets.

The top individual predictors across both cohorts were family history of depression, childhood trauma, female sex, family history of anxiety, and PRS of MDD (**Figure 3E-F**). For MDD severity, childhood trauma was the strongest predictor for the number of episodes in GLAD+ and for age of onset both in GLAD+ and UKB (**Figures S3 and S4**).

The explained variance differed between GLAD+ and UKB by 7–10% points, and the difference in AUC was 0.08–0.10 (**Table 1**). When training on GLAD+ and testing on UKB, both the explained variance and AUC were the same as when training on UKB, and vice versa, indicating good generalizability.

**Table 1.** The replication of MDD and MDD episode prediction across GLAD+ and UKB.

| Phenotype | Cohort | GLAD+ | UKB | UKB<br>resample | UKB | GLAD+ |
| --- | --- | --- | --- | --- | --- | --- |
|  | Dataset | Training | Testing | Testing | Training | Testing |
| MDD | Beta (SE) | 0.55<br>(0.007) | 0.41<br>(0.003) | 0.45<br>(0.018) | 0.41<br>(0.003) | 0.52<br>(0.007) |
|  | R<sup>2</sup> | 33% | 23% | 28% | 23% | 30% |
|  | AUC | 0.84 | 0.74 | 0.78 | 0.74 | 0.82 |
| MDD<br>episodes | Beta (SE) | 0.51<br>(0.008) | 0.33<br>(0.003) | 0.38<br>(0.019) | 0.35<br>(0.003) | 0.50<br>(0.008) |
|  | R<sup>2</sup> | 27% | 10% | 15% | 12% | 25% |

Note: Beta represents the effect size of the prediction scores generated by the model, which uses all predictors in the model to predict MDD. UKB resample, involves randomly selecting a total of 2,442 (1,597 cases and 845 controls) participants across different episodes, mapping to the distribution of MDD episodes in the GLAD+ study.

We counted the number of MDD episodes as a measure of severity in both datasets. The GLAD+ trained model explained 27% of the variance in MDD episodes, when tested in the UKB sample, it explained 10% (**Table 1**). After resampling UKB data to match GLAD+’s MDD severity distribution (**Figure 2**), the explained variance increased to 15% with the model tested in the resampled UKB sample, compared to 12% from the model trained on and then applied in the UKB sample. Additionally, when trained on UKB and applied in GLAD+, the model explained 25% of the variance in MDD episodes, close to the 27% from the model trained in GLAD+.

Similar patterns were found for the age of onset, with predictors explaining 9% of the variance in the GLAD+ trained model and 6% when tested in UKB (**Table S7**). Conversely, the UKB trained model explained 7% of the variance and 8% when tested in GLAD+.

We also applied the model trained for MDD diagnosis to other outcomes (i.e., number of episodes, age of onset, and the degree of treatment resistance, **Table S8**). In GLAD+, the model trained for MDD explained 27% of the variance of the number of episodes, 9% of age of onset, and 0.5% of the degree of treatment resistance (p=1.44×10^−6^). In the UKB, it explained 11% of the variance of the number of episodes and 6% of age of onset. This demonstrates that predictors of MDD diagnosis also predict severity well but do not predict treatment resistance as effectively.

The prediction model assigns each participant a likelihood of being an MDD case. **Figure 4** shows that a higher prediction score is associated with more MDD episodes in both samples. GLAD+ participants have higher prediction scores than UKB participants, reflecting the greater proportion of severe MDD cases in GLAD+ than in UKB.

**Figure 4.**
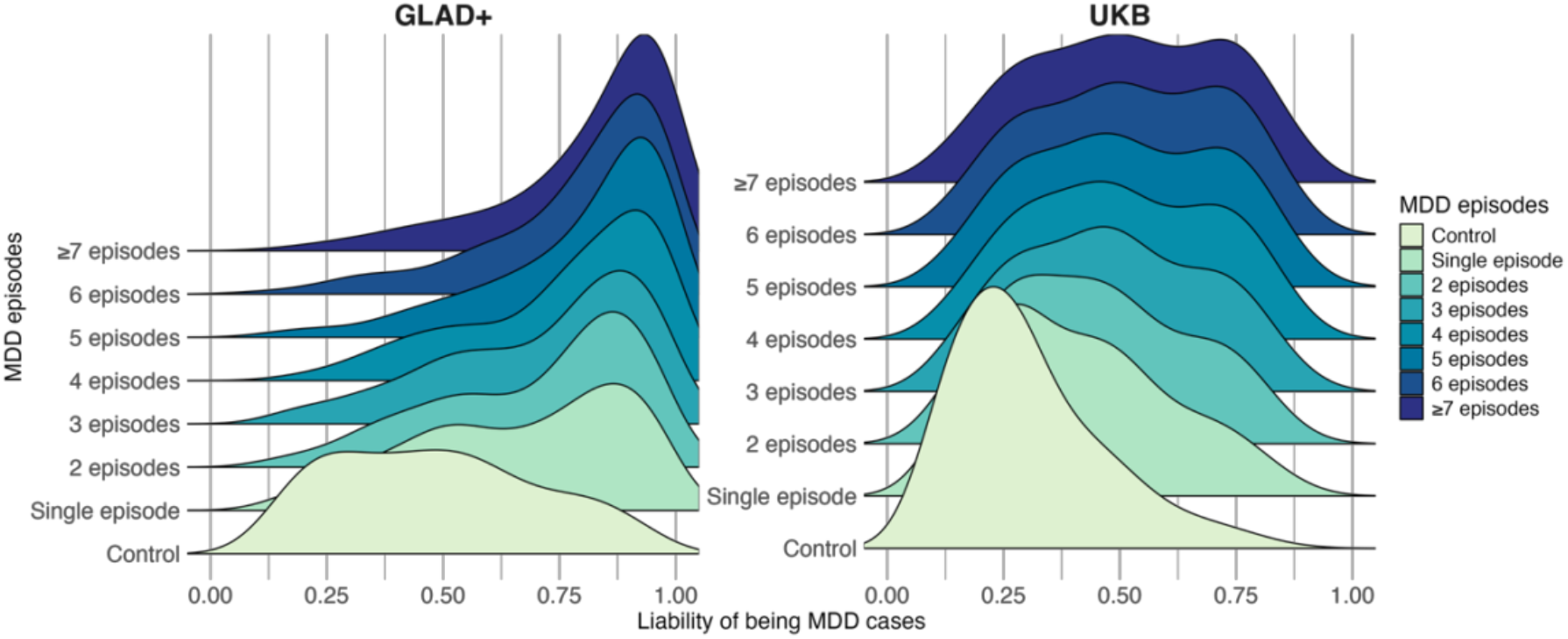
The distribution of the prediction score across frequency of MDD episodes in GLAD+ (n=12,876) and UKB (n=102,821)

Using a 0.5 cut-off, the GLAD+ external validation test model shows high positive predictive value (PPV=0.86), with sensitivity of 0.77 and specificity of 0.72, but low negative predictive value (NPV= 0.58) (**Table S9**). The UKB external validation test model shows high NPV (0.76) and specificity (0.79), but lower PPV (0.60) and sensitivity (0.56) (**Table S9**). Similar patterns appeared in the GLAD+ and UKB trained models (**Table S10**). Calibration plots showing model fitness are in **Figure S7** and **Table S11**. The four predictor groups are interrelated (correlations: 0.15-0.17, **Table S12**) due to shared genetics, environment, and gene-environment interactions.

## Discussion

Using a prediction model, we explained up to 33% of the variance in MDD diagnosis and achieved AUCs of up to 0.84 in differentiating MDD cases from controls in two large UK samples: the GLAD+ Study and the UKB. The strongest individual predictors are family history of depression and childhood trauma. Furthermore, our model also predicted the number of MDD episodes, and age of onset, highlighting its potential for assessing depression severity and identifying individuals at risk of recurrence, and pending clinical validations.

Both family history and PRS of multiple psychiatric disorders and relevant traits explained more variance in MDD than single-family history and single PRS. In the iPSYCH dataset, combining 937 PRSs tripled the explained variance for affective disorder^55^. Similarly, in our GLAD+ study, including family history of 22 psychiatric disorders explained 17% of the variance, compared to 14% using only family history of depression. Incorporating PRSs of multiple traits further increased MDD prediction accuracy, with R^2^ improving by up to 2%.

Family history of psychiatric disorders explained two-to-four times more variance (17% in GLAD+ and 13% in UKB) than PRS of multiple traits (7% and 4%). PRS captures only common genetic variants from GWASs that have not yet reached saturated power^56^, while family history may capture rare and non-additive genetic variants, shared family environment, and learned behaviour or psychosocial factors^57^. As GWASs improve, PRS-explained variance will increase and approach SNP-based heritability^56,58^. Incorporating rare variants may further improve predictions, as seen in a study of BMI, where including rare variants increased SNP-based heritability from 0.24 to 0.29^59^. Whole-genome sequence data may help to identify the remaining common genetic variants and rare variants as currently captured by family history. Combining family history and mPRS improved MDD prediction, with mPRS adding 2-5% variance explained (Figure S1), consistent with a UKB study, where a PRS explained 0.9%, family history 6.1%, and their combination 6.7% for depression^60^.

Family history of depression is the strongest predictor of MDD, but childhood trauma best predicts the number of episodes and age of onset. Consistent with previous findings, individuals reporting a family history of depression are 2-3-times more likely to develop MDD^25^, while childhood trauma increases the risk of earlier onset and treatment-resistant depression by 3-4-times^29^. Notably, while PRSs are not yet widely available in clinical settings, our model still demonstrated strong predictive performance without PRS predictors (ROC-AUC: 0.73-0.82, Table S13). Before PRSs reach their saturated power^56^ and incorporate rare and non-additive genetic variants, assessing family history and childhood trauma remains a practical and robust approach for routine clinical evaluations, given their strong predictive value and ease of measurement.

We replicated our prediction model across the GLAD+ and UKB datasets. The model was trained for MDD diagnosis, MDD episodes, and age of onset in one dataset and tested in the other. The difference in variance explained by the model across the two datasets (7–10%) is likely due to GLAD+ having more severe MDD cases and UKB having milder MDD cases. However, the calibration plots indicated that the model trained in GLAD+ is overconfident, while the model trained in UKB is underconfident. This is mainly due to the imbalanced case-control sample, as GLAD+ has more severe MDD cases. In the resampled UKB test dataset, which matched the MDD severity distribution in GLAD+, the model explained more variance in MDD and its episodes than when trained solely on UKB. Prediction scores for MDD liability showed higher accuracy for positive cases in GLAD+ and negative controls in UKB, likely due to the different proportions of MDD cases across the two datasets (69% vs. 36%). Additionally, our MDD-trained model significantly predicted treatment-resistant depression in GLAD+, but with low variance explained (0.5%). However, caution is warranted in interpreting these results, as further model development and validation is needed before clinical implementation^61^.

Our study has several limitations. First, we only included participants of European genetic ancestry, limiting generalizability to other genetic ancestries. Second, in GLAD+, controls were from the COPING study, where participants were on average 13 years older than those in GLAD (78% MDD cases). This age difference may reflect cohort effects, and age was not included as a predictor. Additionally, the UKB cohort consists of older participants (mean age 56), which may introduce recall bias in reporting the age of onset for MDD. Third, GLAD+ and UKB have unbalanced cases and controls, with GLAD+ having more severe MDD cases. We addressed this using class weights and by resampling UKB to match the severe MDD case distribution in GLAD+. Additionally, family history is a subjective, self-reported measure, which carries a risk of misreporting and may introduce recall bias and ascertainment bias. Finally, family history is an optional question in GLAD+, completed by a subset of participants. Those with affected family members may have been more likely to consider the survey relevant and choose to participate, potentially introducing selection bias.

In this study, we present the first predictive model for depression diagnosis, severity and treatment resistance that incorporates a range of easily accessible predictors. By integrating demographics, psychosocial, clinical and genetic factors, our model explains a substantial proportion of outcome variance, offering a robust tool for early identification and risk stratification. This approach has significant implications for preventative strategies, as it enables targeted interventions in at-risk populations before the onset or escalation of depressive symptoms. Future research should focus on validating this model in diverse cohorts and refining its predictive accuracy through longitudinal studies, ultimately contributing to more personalized and effective mental health care.

## Data and code availability

The GLAD+ Study data are available via a data access request application to the NIHR BioResource (https://bioresource.nihr.ac.uk/using-our-bioresource/academic-and-clinical-researchers/apply-for-bioresource-data/).

The code for defining lifetime depression in MHQ2 in UKB is available: https://github.com/ColemanResearchGroup/MHQ2.

## Acknowledgements

We thank the GLAD+ Study, NIHR BioResource, and UK Biobank volunteers for their participation and gratefully acknowledge the contributions of NIHR BioResource centres, NHS Trusts, UK Biobank, and staff. We also thank the National Institute for Health & Care Research (NIHR), NHS Blood and Transplant, and Health Data Research UK as part of the Digital Innovation Hub Programme.

This study represents independent research funded by the NIHR Biomedical Research Centre at South London and Maudsley NHS Foundation Trust and King’s College London. Further information is available at https://www.maudsleybrc.nihr.ac.uk/facilities/bioresource/.

The views expressed are those of the authors and do not necessarily reflect those of the NHS, NIHR, HSC R&D Division, King’s College London, or the Department of Health and Social Care.

