## Supplementary for "Combined family history and polygenic score prediction of major depressive disorder"

#### Supplementary materials

Rujia Wang[1,2], PhD; Helena L. Davies[1,3,4], PhD; Sang-Hyuck Lee[1,2], MSc; Johan Zvrskovec[1,2], PhD; Christopher Hübel[1,2,5], MD, PhD; Saakshi Kakar[1,2], MSc; Chelsea Malouf[1,2], MSc; Laura Meldrum[1,2], BSc; Yuhao Lin[1,2], BSc; Iona Smith[1,2], MSc; Gursharan Kalsi[1,2], PhD; Henry C. Rogers[1,2,6], MSc; Shannon Bristow[1,2], MSc; Molly R. Davies[7], PhD; Donald M. Lyall[8], PhD; Allan H. Young[9], PhD; Anthony J. Cleare[10,11], PhD; Katrina A. S. Davis[7], MD; Roland Zahn[9], MD, PhD; Victor A. Gault[12], PhD; Le Roy C. Dowey[12], PhD; Ruth K. Price[12], PhD; Keith G. Thomas[12], PhD; James T. R. Walters[13], PhD; Daniel J. Smith[14], PhD; Chérie Armour[15], PhD; Ian R. Jones[16], PhD; Nathalie Kingston[17,18], PhD; John R. Bradley[17], PhD; Andrew M. McIntosh[19], MD; the NIHR BioResource consortium, the GLAD Study, Matthew Hotopf[2,7], PhD; Jonathan R.I. Coleman[1,2], PhD; Evangelos Vassos[1,2], MD, PhD; Thalia C. Eley[1,2], PhD; Raquel Iniesta[20,21], PhD; Gerome Breen[1,2]*, PhD

*1. Social, Genetic, and Developmental Psychiatry Centre; Institute of Psychiatry, Psychology and Neuroscience; King's College London, London, UK*

*2. National Institute for Health and Care Research Maudsley Biomedical Research Centre, South London and Maudsley NHS Trust, London, UK*

*3. Mental Health Center Ballerup, Copenhagen University Hospital – Mental Health Services CPH, Center for Eating and feeding Disorders Research, Copenhagen, Denmark*

*4. Institute of Biological Psychiatry, Mental Health Center Sct. Hans, Mental Health Services Copenhagen, Roskilde, Denmark*

*5. National Centre for Register-based Research, Aarhus University, Aarhus, Denmark*

*6. Department of Psychiatry, Icahn School of Medicine at Mount Sinai, New York, NY, USA*

*7. Department of Psychological Medicine, Institute of Psychiatry, Psychology & Neuroscience, King's College London, London, UK*

*8. School of Health and Wellbeing, University of Glasgow, Glasgow, UK*

*9. South London and Maudsley NHS Foundation Trust and Centre for Affective Disorders, Department of Psychological Medicine, Institute of Psychiatry, Psychology, and Neuroscience, King's College London, London, UK*

*10. The Institute of Psychiatry, Psychology and Neuroscience, King's College London, London, UK*

*11. South London and Maudsley NHS Foundation Trust, Maudsley Hospital, London, UK*

*12. School of Biomedical Sciences, Faculty of Life and Health Sciences, Ulster University, Coleraine, UK*

*13. Centre for Neuropsychiatric Genetics and Genomics, Division of Psychological Medicine and Clinical Neurosciences, School of Medicine, Cardiff University, Cardiff, UK*

*14. Division of Psychiatry, Centre for Clinical Brain Sciences, University of Edinburgh, Royal Edinburgh Hospital, Edinburgh, UK*

*15. Research Centre for Stress Trauma and Related Conditions (STARC), School of Psychology, Queen's University Belfast, Belfast, UK*

*16. National Centre for Mental Health, Cardiff University, Cardiff, UK*

*17. NIHR BioResource, Cambridge University Hospitals NHS Foundation Trust, Cambridge Biomedical Campus, Cambridge, UK*

*18. Department of Haematology, School of Clinical Medicine, University of Cambridge, Cambridge Biomedical Campus, Cambridge, UK*

*19. Division of Psychiatry, Centre for Clinical Brain Sciences, University of Edinburgh, Edinburgh, UK*

*20. Department of Biostatistics and Health Informatics, King's College London, Denmark Hill, Camberwell, London, UK*

*21. King's Institute for Artificial Intelligence, King's College London, London, UK*

** Corresponding author:* Gerome Breen,, Social Genetic and Developmental Psychiatry Centre, King's College London, 16 De Crespigny Park, Denmark Hill, London, United Kingdom, SE5 8AF; Office: +44 207 848 0409; Mobile: +44 7718 120711

### Supplementary Methods

#### Study samples

##### GLAD+ study

The GLAD+ study includes two cohorts: the Genetic Links to Anxiety and Depression Study (GLAD) and the NIHR BioResource COVID-19 Psychiatric and Neurology Genetics study (COPING) in the UK. The GLAD Study (www.gladstudy.org.uk) is an ongoing study that recruits participants who have experienced depression and/or anxiety online (via media campaigns). Participants provide demographic, environmental, and genetic data and consent to medical record linkage and potential recontact. Currently, the GLAD study has over 66,000 consented participants, with more than 50,000 having completed the online survey, and over 36,000 saliva samples collected. During the COVID-19 pandemic, the GLAD Study research team recontacted GLAD participants and healthy volunteers from other NIHR BioResource studies to conduct the COVID-19 Psychiatric and Neurology Genetics study (COPING). The goal was to estimate the impact of the COVID-19 pandemic on mental and neurological health. We recruited over 20,000 participants with common mental disorders and over 11,000 healthy volunteers, two-thirds of whom have been genotyped. All participants provided informed consent for the GLAD and/or COPING studies and the NIHR BioResource (https://bioresource.nihr.ac.uk/).

##### UK Biobank

The UK Biobank (UKB) is a prospective health study of over 500,000 individuals in the UK, aged 40-69 at recruitment (2006-2010). In 2016-17, participants were invited via email and postal newsletter to complete the first online Mental Health Questionnaire (MHQ1), resulting in responses from over 157,000 individuals by July 2017[46]. In 2022, the second Mental Health Questionnaire (MHQ2) was sent out to gather repeat data, new data, and data from participants who had not completed the first questionnaire. This led to responses from over 169,000 individuals in October-November 2022, including over 110,000 participants who had completed the earlier questionnaire (MHQ1 in 2016) and over 52,000 who had not. Consequently, more than 206,000 UK Biobank participants have data for either MHQ1 or MHQ2. Both MHQ1 and MHQ2 measured lifetime depression, anxiety disorders, and other psychiatric disorders. In MHQ2, self-reported family history of psychiatric disorders among first-degree relatives was collected. All participants provided informed consent for the UKB.

##### Major depressive disorder (MDD)

Lifetime major depressive disorder was accessed using the Adapted Composite International Diagnostic Interview-Short Form (CIDI-SF) in GLAD+ and UKB. According to the Diagnostic and Statistical Manual of Mental Disorders Fifth Edition (DSM-5) criteria, MDD cases were defined as individuals who have five or more of the following symptoms, with at least one of the core symptoms occurring daily or nearly every day for a duration of two weeks or more. The two core symptoms include depressed mood and loss of interest. The additional symptoms include sleep difficulties (insomnia or hypersomnia), weight changes, fatigue, diminished ability to think or concentrate, feelings of worthlessness and suicidal ideation. In total, the GLAD+ study included 20,191 MDD cases and 11,064 controls.

Based on the same criteria, we combined depression diagnoses across MHQ1 and MHQ2 in the UKB. Controls were defined as participants who had not self-reported a diagnosis of depression, did not experience the two core symptoms, and/or had a Patient Health Questionnaire-9 (PHQ-9) total score less than 5. Individuals diagnosed with depression in either MHQ1 or MHQ2 were defined as cases, while those without depression in both waves or measured in only one wave without a depression diagnosis were defined as controls. In total, the UKB study included 54,670 MDD cases and 126,381 controls. The CIDI-SF items in MHQ1 and MHQ2 of the UKB are shown in **Table S1**. In both GLAD+ and UKB, we also excluded participants among the controls who reported experiencing episodes of depression.

**Table S1**. CIDI-SF Items in MHQ1 and MHQ2 of the UKB

| MDD (CIDI-SF) | MHQ1 | MHQ2 |
|---|---|---|
| **2 core symptoms** | | |
| depressed mood | 20446 | 29011 |
| loss of interest | 20441 | 29012 |
| **additional symptoms** | | |
| sleep difficulties | 20532 | 29022 |
| weight changes | 20536 | 29021 |
| fatigue | 20449 | 29018 |
| diminished ability to think or concentrate | 20435 | 29026 |
| feelings of worthlessness | 20450 | 29027 |
| suicidal ideation | 20437 | 29029 |

#### Number of MDD episodes

The number of MDD episodes was defined by asking participants the following question: “Please estimate the number of times you have had periods of depression or low mood in your life lasting two or more weeks.” The answers ranged from 1 to 12, with an additional category for 13 or more in GLAD+. UKB asked a similar question about periods of feeling depressed for at least two weeks. In UKB, the index codes for the number of MDD episodes are 20442 in MHQ1 and 29033 in MHQ2.

#### Age of MDD onset

Age of MDD onset was determined by asking participants their age during their first two-week period of depression in both cohorts. In UKB, the index codes for the MDD age of onset are 20433 in MHQ1 and 29034 in MHQ2.

#### Treatment-resistant depression

Treatment-resistant depression (TRD), according to the Maudsley Staging Method (MSM), was assessed using a combination of the 4-item MSM and the depression severity score measured by the PHQ-9 in GLAD+. The PHQ-9 severity scoring categorizes depression as follows: 0–4 as none (0), 5–9 as mild (1), 10–14 as moderate (2), 15–19 as moderately severe (3), and 20–27 as severe (4). These scores can be recoded into a scale of 0–4. Participants were only shown the MSM if they had mild or more severe depression based on the PHQ-9 items, with scores ranging from 0 to 10. The MSM-TRD sum score combined the MSM score and PHQ-9 score, ranging from 0 to 14.

**Table S2.** Maudsley Staging Method four-items

| CODE | Questions | Answers |
|---|---|---|
| MSM-1:<br>when low mood begin | How long ago did your current or most recent episode of depression or low mood begin? | 1= Less than 1 year ago<br>2= 1-2 years ago<br>3= More than 2 years ago |
| MSM-2:<br>taking antidepressants for 6 weeks or longer | During the current or most recent episode of depression or low mood, how many antidepressant medications have you taken for 6 weeks or longer? | 0= None<br>1= One to two<br>2= three to four<br>3= five to six<br>4= seven to ten<br>5= More than 10 |
| MSM-3:<br>add-on medication | If individuals don’t respond fully to antidepressants, doctors sometime prescribe “add-on” or “augmentation” medications in addition to the antidepressant (such as lithium, quetiapine or aripiprazole).During the current or most recent episode of depression or low mood, have you taken an add on medication for 6 weeks or longer? | 0= No<br>1= Yes |

| | | |
|---|---|---|
| MSM-4:<br>ECT | Have you received electroconvulsive therapy (ECT) in this current or most recent episode of depression or low mood? (Please only answer yes if your course of ECT included 8 treatment sessions or more.) | 0= No<br>1= Yes |

#### Family history (FH)

In GLAD+, family history was assessed by asking participants, "Have any of your family members ever been diagnosed with one or more of the following mental health disorders by a professional, even if they don't have it currently?" The mental health disorder options available included depression, depression during or after pregnancy, premenstrual dysphoric disorder (PMDD), bipolar disorder (BD), generalized anxiety disorder (GAD), social anxiety, specific phobia, agoraphobia, panic attacks, panic disorder (PD), post-traumatic stress disorder (PTSD), obsessive-compulsive disorder (OCD), body dysmorphic disorder (BDD), schizophrenia (SCZ), psychosis or psychotic illness, personality disorder, autism spectrum disorder (ASD), attention deficit / hyperactivity disorder (ADHD), anorexia nervosa (AN), atypical anorexia nervosa, bulimia nervosa (BN), and binge-eating disorder (BED) (**Table S3**). In an optional survey of the GLAD study, family history was assessed by asking participants: "How many of your immediate family members (parents, brothers, sisters, or children) have ever been diagnosed with the following mental health disorders?" Only 6,000 participants responded to this question. While COPING and UKB participants were not asked the same question, we used a binary family history variable (yes/no) in the analysis.

In UKB, family history was assessed by asking participants, "Have any of your first degree blood relatives had any of the following conditions?" in MHQ2 (Index code 29001). The condition options available included depression, BD, anxiety disorder, SCZ, psychosis, personality disorder, ASD, ADHD, and eating disorder (ED) (**Table S4**). Participants who responded to the provided options for specified mental health disorders were considered to have a family history of corresponding mental health disorders. For the external dataset validation, family history of psychiatric disorders in GLAD+ were mapped the same as in UKB.

**Table S3**. Characteristics of 22 family history predictors in GLAD+

| **Family history** (FH, yes, %) | **Total** (n=14,379) | **MDD cases** (n=9,927) | **Controls** (n=4,452) |
|---|---|---|---|
| depression | 7,116 (49.5) | 6,119 (61.6) | 997 (22.4) |
| depression during or after pregnancy | 1,440 (10.0) | 1,309 (13.2) | 131 (2.9) |
| premenstrual dysphoric disorder | 92 (0.64) | 88 (0.89) | 4 (0.09) |
| bipolar disorder | 1,219 (8.5) | 1,094 (11.0) | 125 (2.8) |

| | | | |
|---|---|---|---|
| generalized anxiety disorder | 4,139 (28.8) | 3,612 (36.4) | 527 (11.8) |
| social anxiety | 935 (6.5) | 868 (8.7) | 67 (1.5) |
| agoraphobia | 356 (2.5) | 333 (33.5) | 23 (0.52) |
| specific phobia | 321 (2.2) | 297 (3.0) | 24 (0.54) |
| panic attacks | 1,626 (11.3) | 1,441 (14.5) | 185 (4.2) |
| panic disorder | 608 (4.2) | 567 (5.7) | 41 (0.92) |
| PTSD | 696 (4.8) | 650 (6.5) | 46 (1.0) |
| SCZ | 596 (4.1) | 528 (5.3) | 68 (1.5) |
| ADHD | 647 (4.5) | 572 (5.8) | 75 (1.7) |
| ASD | 1,177 (8.2) | 1,021 (10.3) | 156 (3.5) |
| OCD | 644 (4.5) | 581 (5.9) | 63 (1.4) |
| BDD | 174 (1.2) | 164 (1.7) | 10 (0.22) |
| Psychosis | 584 (4.1) | 525 (5.3) | 59 (1.3) |
| Personality disorder | 475 (3.3) | 443 (4.5) | 32 (0.72) |
| Anorexia nervosa | 581 (4.0) | 498 (5.0) | 83 (1.9) |
| Atypical anorexia nervosa | 36 (0.25) | 33 (0.33) | 3 (0.07) |
| Bulimia nervosa | 301 (2.1) | 267 (2.7) | 34 (0.76) |
| Binge-eating disorder | 139 (0.97) | 130 (1.3) | 9 (0.20) |

Abbreviation: PTSD, post-traumatic stress disorder; OCD, obsessive-compulsive disorder; BDD, body dysmorphic disorder; SCZ, schizophrenia; ASD, autism spectrum disorder; ADHD, attention deficit / hyperactivity disorder.

**Table S4**. Characteristics of mapped predictors in GLAD+ and UKB

| **Characteristics** | **GLAD+** | | | **UKB** | | |
|---|---|---|---|---|---|---|
| | Total (n=14,379) | MDD cases (n=9,927) | Controls (n=4,452) | Total (n=111,422) | MDD cases (n=40,667) | Controls (n=70,755) |
| Age (mean±sd) | 48.4 ± 15.6 | 44.3 ± 14.7 | 57.5 ± 13.5 | 56.0 ± 7.5 | 54.2 ± 7.4 | 57.0 ± 7.4 |
| Sex (female, %) | 10,179 (70.8) | 7,945 (80.0) | 2,234 (50.3) | 62,452 (56.0) | 28,457 (70.0) | 33,995 (48.0) |
| **Family history** (yes, %) | | | | | | |
| depression | 7,116 (49.5) | 6,119 (61.6) | 997 (22.4) | 28,373 (25.5) | 17,352 (42.7) | 11,021 (15.6) |
| anxiety | 4,552 (31.7) | 3,960 (39.9) | 592 (13.3) | 11,248 (10.1) | 6,683 (16.4) | 4,565 (6.5) |
| bipolar disorder | 1,219 (8.5) | 1,094 (11) | 125 (2.8) | 3,274 (2.9) | 1,928 (4.7) | 1,346 (1.9) |
| eating disorder | 925 (6.4) | 804 (8.1) | 121 (2.7) | 3,248 (2.9) | 1,811 (4.5) | 1,437 (2.0) |
| SCZ | 596 (4.1) | 528 (5.3) | 68 (1.5) | 1,685 (1.5) | 893 (2.2) | 792 (1.1) |
| personality disorder | 475 (3.3) | 443 (4.5) | 32 (0.72) | 1,398 (1.3) | 940 (2.3) | 458 (0.64) |
| psychosis | 584 (4.1) | 525 (5.3) | 59 (1.3) | 1,679 (1.5) | 967 (2.4) | 712 (1.0) |
| ADHD | 647 (4.5) | 572 (5.8) | 75 (1.7) | 2,125 (1.9) | 1,260 (3.1) | 865 (1.2) |
| ASD | 1,177 (8.2) | 1,021 (10.3) | 156 (3.5) | 3,315 (3.0) | 1,915 (4.7) | 1,400 (2.0) |

| | | | | | | |
|---|---|---|---|---|---|---|
| **Childhood trauma** (yes, %) | 5,754 (40.0) | 5,020 (50.6) | 734 (16.5) | 31,051 (27.9) | 17,019 (41.8) | 14,032 (19.8) |
| MDD PRS (mean±sd) | 0 ± 1 | 0.16 ± 0.97 | -0.36 ± 0.98 | 0 ± 1 | 0 ± 1 | 0 ± 1 |

Abbreviation: MDD, major depressive disorder. PRS, polygenic risk score. SD, standard deviation.

#### Childhood trauma (ChT)

Reported childhood trauma was assessed using the Childhood Trauma Survey[50], which includes five items measured on a Likert scale. Each item pertains to the frequency of a specific type of abuse (emotional, physical, sexual) or neglect (emotional, physical). Participants who reported experiencing at least one of these five types of childhood trauma were classified as individuals who had reported childhood trauma. Details of the five items assessing childhood trauma are shown in **Table S5**.

**Table S5**. Items and definitions of childhood trauma

| Childhood trauma | Questions | Definition | MHQ1&MHQ2 |
|---|---|---|---|
| Emotional abuse | I felt that someone in my family hated me | If the answer is "Never true" or "Rarely true", defined as no emotional abuse;<br><br>If the answer is "Sometimes true" or "Often true" or "Very often true", it is defined as having emotional abuse. | 20487 & 29078 |
| Emotional neglect | When I was growing up, I felt loved | If the answer is "Sometimes true" or "Often true" or "Very often true", defined as no emotional neglect;<br><br>If the answer is "Never true" or "Rarely true", it is defined as having emotional neglect. | 20489 & 29076 |
| Physical abuse | People in my family hit me so hard that it left me with bruises or marks | If the answer is "Never true" or "Rarely true", defined as no physical abuse; | 20488 & 29077 |

| | | | |
|---|---|---|---|
| | | If the answer is “Sometimes true” or “Often true” or “Very often true”, it is defined as having physical abuse. | |
| Physical neglect | There was someone to take me to the doctor if I needed it | If the answer is “Sometimes true” or “Often true” or “Very often true”, defined as no physical neglect;<br><br>If the answer is “Never true” or “Rarely true”, it is defined as having physical neglect. | 20491 & 29080 |
| Sexual abuse | Someone molested me (sexually) | If the answer is “Never true”, defined as no sexual abuse;<br><br>If the answer is “Rarely true” or “Sometimes true” or “Often true” or “Very often true”, it is defined as having sexual abuse. | 20490 & 29079 |

#### Genetic data in GLAD+ and UKB

##### GLAD+ study

All data from GLAD+ study were genotyped by ThermoFisher on the UK Biobank Axiom Array v1 and v2 across numerous genotyping batches. Genetic data for GLAD+ study were restricted to individuals from European ancestries (749,044 SNPs before quality control). Ancestry was determined using GenoPred ( https://opain.github.io/GenoPred/index.html), by projecting GLAD+ individuals on genomic principal components from the 1000 Genomes reference data, and assigning individuals a genetic ancestry if they lay < 3 SD from the mean of individuals from that ancestry superpopulation in 1000 Genomes. Quality control was conducted, excluding variants with MAF < 0.01, call rate < 0.95, or which were deviant from Hardy-Weinberg equilibrium (p < 10^-10). Individuals were excluded if they had withdrawn from the study following genotyping, if they were a duplicate of a higher-quality sample (not including known identical twins), if they were known to be mislabelled, if their genotypic sex (males Fx > 0.8, females Fx < 0.5) did not match their sex assigned at birth, if they were outliers on genome-wide heterozygosity (absolute(Fhat) > 0.2), or if they had an excess of relatives (average pi-hat > 3 SD from the mean). Following quality control, 33,635 individuals and 484,182 variants were available for imputation. Imputation was carried out to TopMED Freeze 8, using the dedicated imputation server ( https://imputation.biodatacatalyst.nhlbi.nih.gov/#!). Following imputation, data was further restricted to data with MAF >= 0.01 and R2 >= 0.3, leaving 15,009,228 variants for analysis.

##### UKB Study

The UKB genetic data contain genotypes for 488,377 participants. We used 461,719 participants of European ancestry to calculate PRSs. Details of genotype arrays and quality controls have been described elsewhere[62].

#### MegaPRS

Polygenic risk scores were generated using genomic data from the GLAD+ study and summary statistics of recent large GWASs for 22 psychiatric disorders (i.e., MDD, anxiety, ASD, ADHD, AN, BD, PTSD, PD, OCD, SCZ, and neuroticism) and related traits (e.g., educational attainment, alcohol dependence, ever smoking, heavy smoking, cannabis use, tiredness, insomnia, subjective well-being, household income, social deprivation and body mass index) (for details, see **Table S6**). In UKB, PRS was generated using genomic data of participants of European ancestry and summary statistics of recent large GWASs without UK/UKB samples for 7 psychiatric disorders and related traits (i.e., MDD, anxiety, ASD, ADHD, BD, PTSD, and BMI). The GenoPred pipeline[51] was utilized, which uses MegaPRS[52] for calculation of PRS. MegaPRS implements polygenic scoring approaches using the LDAK heritability model, where the variance explained by each SNP depends on its allele frequency, linkage disequilibrium and functional annotations. The LDAK model was first trained on genetic data from 90% of the participants and the remaining 10% of the participants were used to select the best-fitting model that maximizes the prediction power of the model. The best-fitting model was then employed to compute MegaPRS for all the participants.

**Table S6**. Sumstats for psychiatric disorders and related traits

| Disorders /traits | Sumstats | PMID/ doi | N |
|---|---|---|---|
| Depression * | MDD PGC3 without UK sample 2024 | 39814019 | Cases: 347,453; Controls: 1,241,648 |
| Anxiety * | Anxiety PGC2 without UKB 2024 | https://doi.org/10.1101/2024.07.03.24309466 | Cases: 94,930; Controls: 655,768 |
| Bipolar disorder * | Bipolar disorder without UKB sample 2021 | 34002096 | Cases: 41,917; Controls: 371,549; - UKB sample |
| Neuroticism | NEURO 2018 | 29500382 | 380,506 |
| Schizophrenia | SCZ 2022 | 35396580 | Cases: 53,386; Controls: 77,258 |
| ADHD * | PGC + iPSYCH 2023 | 36702997 | Cases: 38,691; Controls: 186,843 |
| ASD * | PGC + iPSYCH | 30804558 | Cases: 18,381; Controls: 27,969 |
| PTSD * | PTSD MVP 2021 | 33510476 | Cases: 36,301; Controls: 178,107 |
| Panic disorder | Panic 2019 | 31712720 | Cases: 2,147; Controls: 7,760 |
| AN | AN PGC2 | 31308545 | Cases: 16,992; Controls: 55,525 |
| OCD | PGC 2017 | 28761083 | Cases: 2,688; Control: 7,037 |
| BMI * | GAINT 2015 | 25673413 | 322,154 |
| Educational attainment | EA 2022 | 35361970 | 3,037,499 |
| Alcohol dependence | Alcohol dependence 2017 | 30482948 | Cases: 11,569; Controls: 34,999 |
| Ever smoking | GSCAN-23andMe 2019 | 30643251 | 632,802 |
| Heavy smoking | UKB 2017 | 28166213 | 46,758 |
| Cannabis use | PGC 2020 | 33096046 | Cases: 17,068; Controls: 357,219 |
| Tiredness | UKB 2017 | 28322280 | 108,976 |
| Insomnia | Insomnia 2019 | 30804565 | 1,331,010 |

| | | | |
|---|---|---|---|
| Subjective well-being | SSGAC 2016 | 27089181 | 298,420 |
| Household income | UKB 2016 | 27818178 | 112,151 |
| Social deprivation | UKB 2016 | 27818178 | 112,151 |

* These sumstats were used for PRSs in both GLAD+ and UKB.

**Prediction model**

**Class weighting for imbalanced sample**

We applied class weighting to address the imbalanced samples in GLAD+ and UKB. In GLAD+, we included 9,927 cases and 4,452 controls (a ratio of 2.23 to 1); thus, we weighted the control group as 2.23 and the case group as 1 in the model. Similarly, in UKB, we included 40,667 cases and 70,755 controls (a ratio of 1 to 1.74); therefore, we weighted the case group as 1.74 and the control group as 1 in the model.

#### Supplementary Results

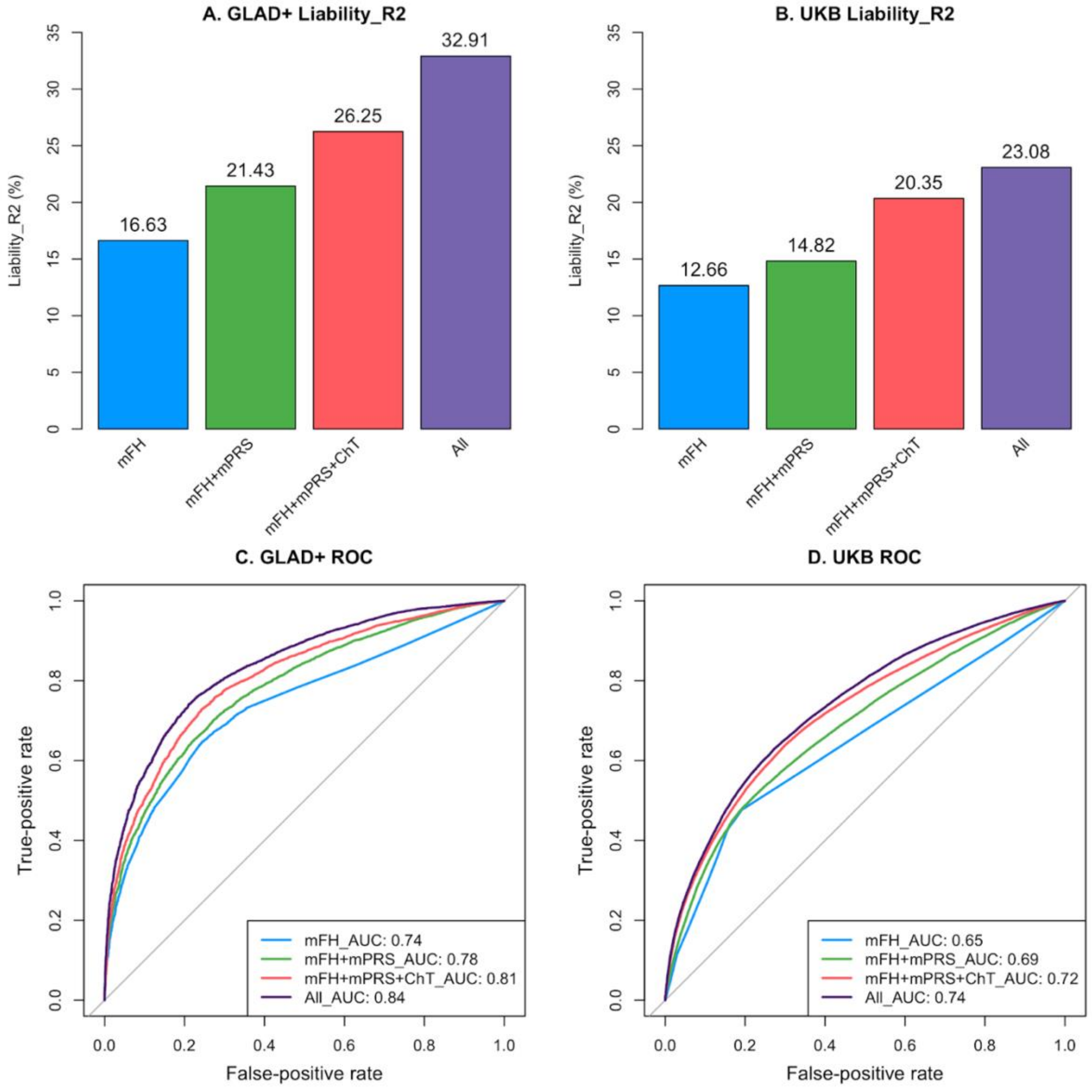


**Figure S1.** The effect of multi-component predictors in GLAD+ and UKB

The liability $R^2$ of MDD explained by a group of predictors in GLAD+ (**Figure S1A**), with a prevalence of MDD of 5%, and in UKB (**Figure S1B**), with a prevalence of MDD of 15%. The ROC of a group of predictors in GLAD+ (**Figure S1C**) and in UKB (**Figure S1D**). mFH, family history of multiple psychiatric disorders, including family history of 22 psychiatric disorders in GLAD+, and family history of 9 psychiatric disorders in UKB. mPRS, polygenic risk scores of multiple traits, including PRS of 11 psychiatric disorders and 11 related traits in GLAD+, and 7 PRSs of psychiatric disorders and BMI based without UK/UKB GWAS sumstats in UKB. ChT, childhood trauma. Demo, sociodemographic factors, including sex and 10 principal components. All, including all groups of predictors. ROC, receiver operating characteristic curve.

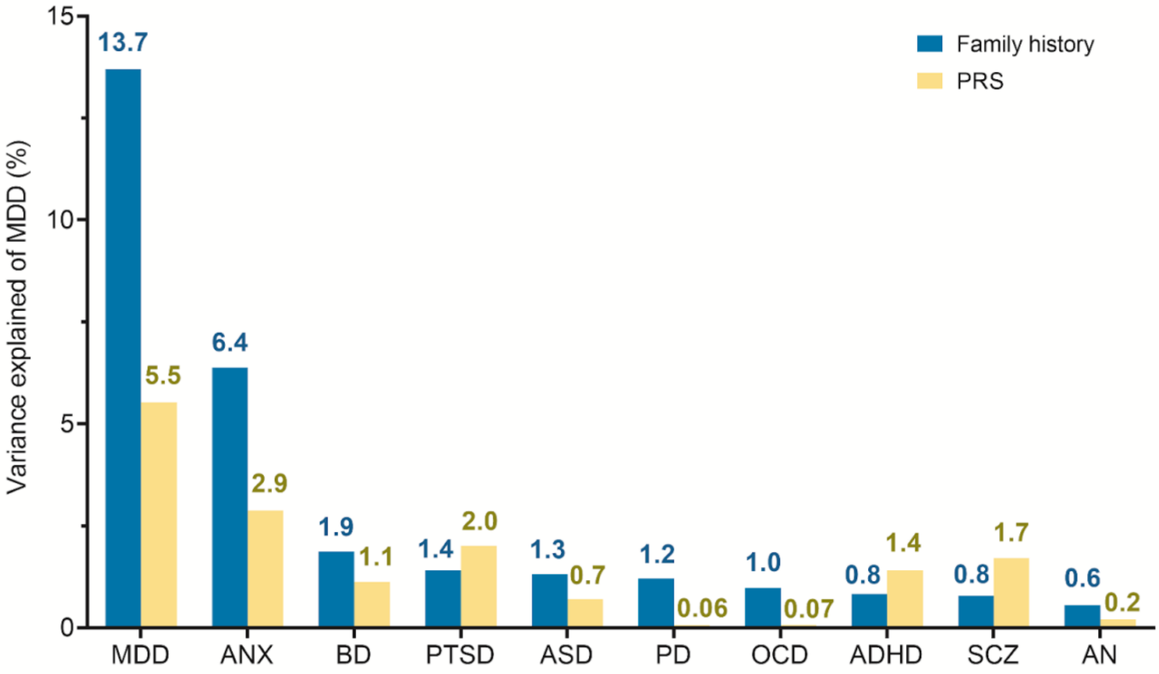


**Figure S2**. Proportion of MDD variance explained by family history and PRS in univariate regression in GLAD+ study

In univariate analysis, family history of psychiatric disorders explained more variance than PRS for MDD, except for PTSD, ADHD and SCZ (Figure S1). Specifically, family history of depression explained 13.7% of the variance for MDD, while PRS explained 5.5% of the variance.

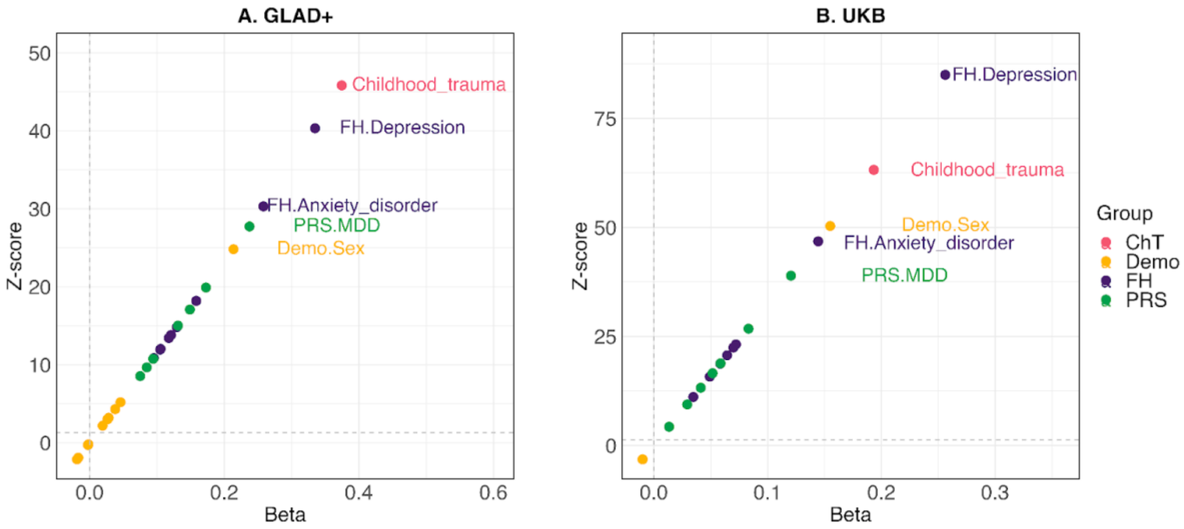


**Figure S3**. The effect of individual predictors for MDD episodes in GLAD+ and UKB
FH, family history, including family history of 9 psychiatric disorders in GLAD+ and UKB. PRS, polygenic risk scores, including 7 PRSs of psychiatric disorders and BMI based without UK/UKB GWAS sumstats in GLAD+ and UKB. ChT, childhood trauma. Demo, sociodemographic factors, including sex and 10 principal

components.

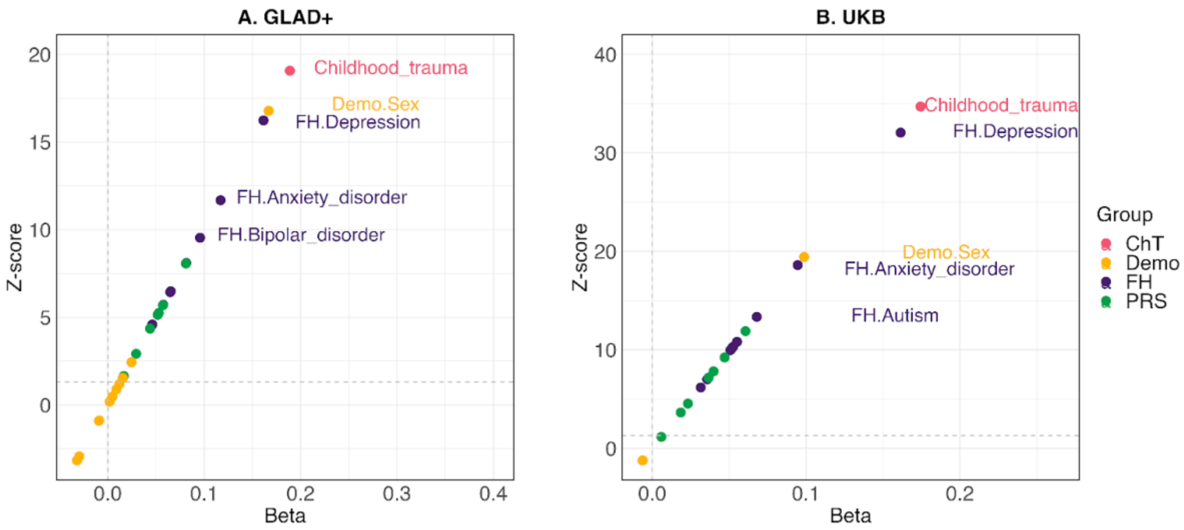


**Figure S4**. The effect of individual predictors for MDD age of onset in GLAD+ and UKB
FH, family history, including family history of 9 psychiatric disorders in GLAD+ and UKB. PRS, polygenic risk scores, including 7 PRSs of psychiatric disorders and BMI based without UK/UKB GWAS sumstats in GLAD+ and UKB. ChT, childhood trauma. Demo, sociodemographic factors, including sex and 10 principal components.

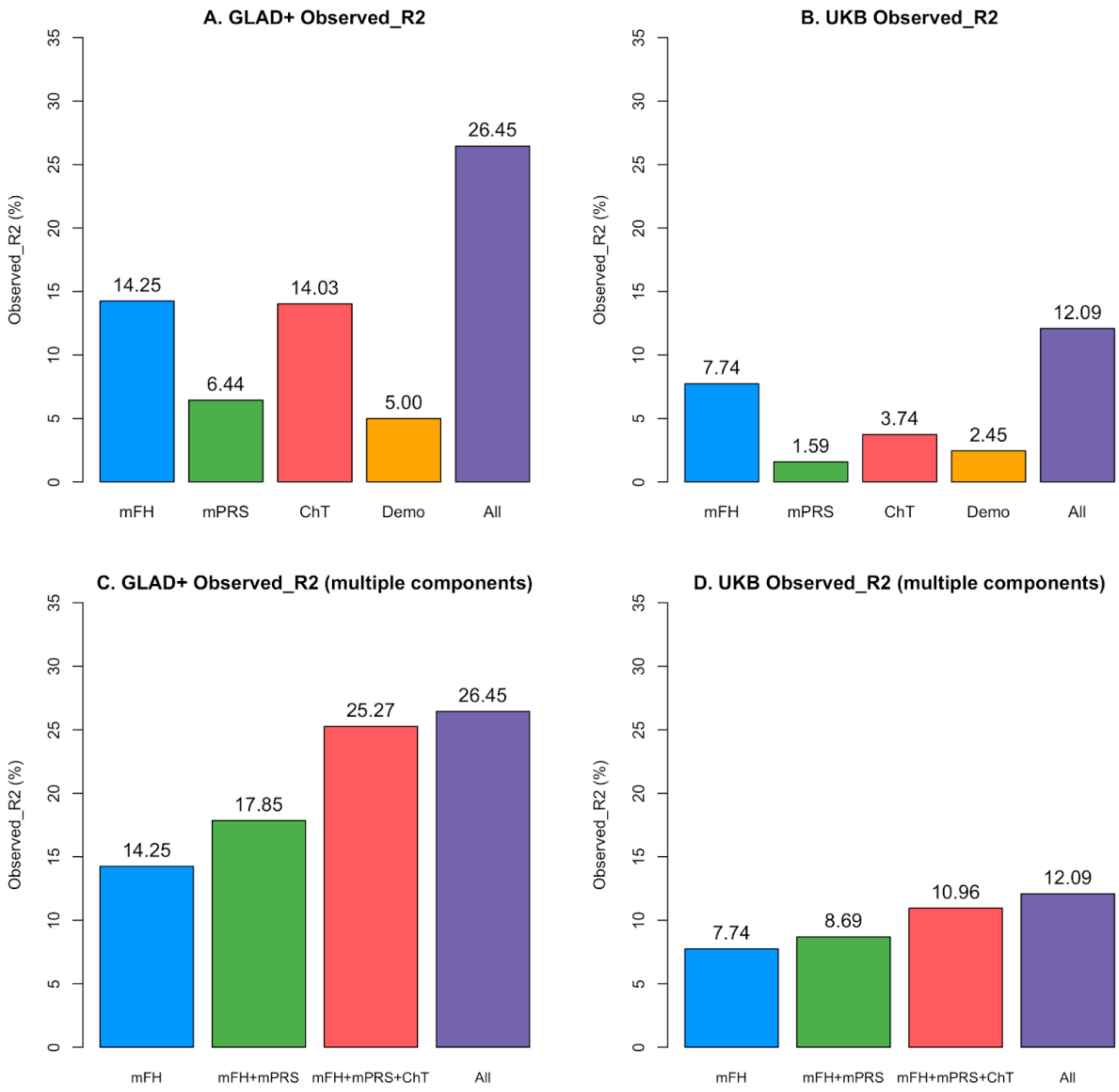


**Figure S5**. Effects of group of predictors for MDD episodes in GLAD+ and UKB

The observed $R^2$ of MDD episodes explained by a group of predictors in GLAD+ (**Figure S5A**), and in UKB (**Figure S5B**). The observed $R^2$ of MDD episodes explained by multi-steps of including groups of predictors in GLAD+ (**Figure S5C**) and in UKB (**Figure S5D**). mFH, family history of multiple psychiatric disorders, including family history of 9 psychiatric disorders in GLAD+ and UKB. mPRS, multi-polygenic risk scores, including 7 PRSs of psychiatric disorders and BMI based without UK/UKB GWAS sumstats in GLAD+ and UKB. ChT, childhood trauma. Demo, sociodemographic factors, including sex and 10 principal components. All, including all groups of predictors.

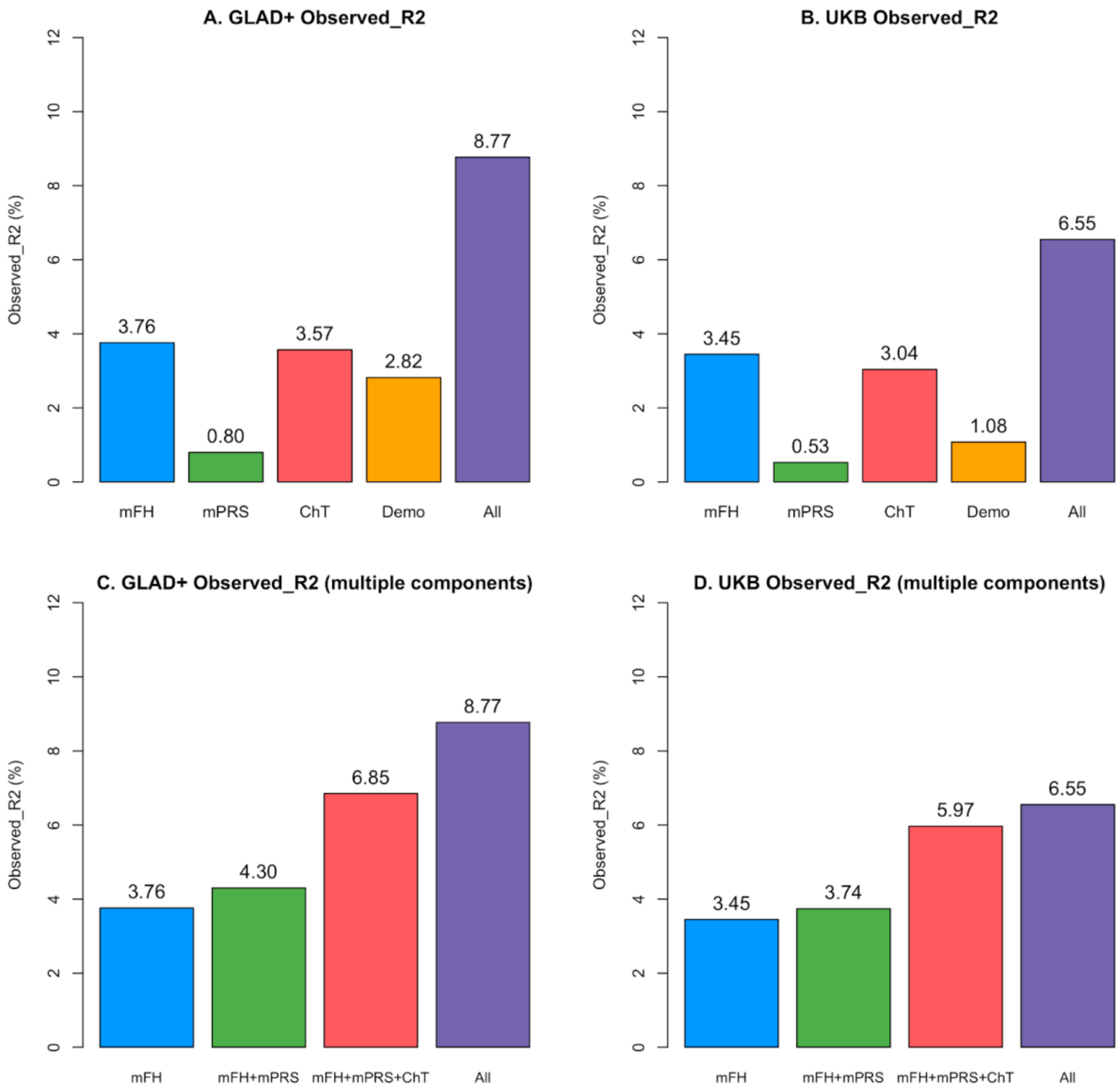


**Figure S6**. Effects of each group of predictors for MDD age of onset in GLAD+ and UKB

The observed $R^2$ of age of onset for MDD explained by a group of predictors in GLAD+ (**Figure S6A**) and in UKB (**Figure S6B**). The observed $R^2$ of age of onset for MDD explained by multi-steps of including groups of predictors in GLAD+ (**Figure S6C**) and in UKB (**Figure S6D**). mFH, family history of multiple psychiatric disorders, including family history of 9 psychiatric disorders in GLAD+ and UKB. mPRS, polygenic risk scores of multiple traits, including 7 PRSs of psychiatric disorders and BMI-based without UK/UKB GWAS sumstats in GLAD+ and UKB. ChT, childhood trauma. Demo, sociodemographic factors, including sex and 10 principal components. All, including all groups of predictors.

**Table S7.** The replication of MDD and MDD episodes, age of onset across GLAD+ and UKB

| Phenotype | Dataset | Cohort | Beta (SE) | $R^2$ (%) | AUC | N |
|---|---|---|---|---|---|---|
| **MDD** | Training | GLAD+ | 0.55 (0.007) | 32.9 | 0.84 | 14,379 |
| | Testing | UKB | 0.41 (0.003) | 23.2 | 0.74 | 111,422 |
| | Testing | UKB-resampled | 0.45 (0.018) | 27.9 | 0.78 | 2,442 |
| | Training | UKB | 0.41 (0.003) | 23.1 | 0.74 | 111,422 |
| | Testing | GLAD+ | 0.52 (0.007) | 29.8 | 0.82 | 14,379 |
| **MDD episodes** | Training | GLAD+ | 0.51 (0.008) | 26.5 | | 12,876 |
| | Testing | UKB | 0.33 (0.003) | 10.1 | | 102,821 |
| | Testing | UKB-resampled | 0.38 (0.019) | 14.5 | | 2,442 |
| | Training | UKB | 0.35 (0.003) | 12.1 | | 102,821 |
| | Testing | GLAD+ | 0.50 (0.008) | 24.8 | | 12,876 |
| **MDD age of onset** | Training | GLAD+ | 0.30 (0.010) | 8.8 | | 9,837 |
| | Testing | UKB | 0.24 (0.005) | 6.0 | | 38,395 |
| | Testing | UKB-resampled | 0.24 (0.025) | 5.5 | | 2,442 |
| | Training | UKB | 0.26 (0.005) | 6.6 | | 38,395 |
| | Testing | GLAD+ | 0.29 (0.010) | 8.2 | | 9,837 |

The UKB resample involves randomly selecting a total of 2,442 (1,597 cases and 845 controls) participants across different episodes, mapping to the distribution of MDD episodes in the GLAD+ study.

**Table S8.** Refit the prediction model for MDD to other phenotypes in GLAD+ and UKB

| Cohort | Dataset | Phenotype | Beta (SE) | $R^2$ (%) | N |
|---|---|---|---|---|---|
| **GLAD+** | Training | MDD | 0.55 (0.007) | 32.9 | 14,379 |
| | Testing | MDD episodes | 0.52 (0.008) | 26.8 | 12,876 |
| | Testing | MDD age of onset | -0.29 (0.010) | 8.6 | 9,837 |
| | Testing | MSM-TRD | 0.07 (0.015) | 0.5 | 4,206 |
| **UKB** | Training | MDD | 0.41 (0.003) | 23.0 | 111,422 |
| | Testing | MDD episodes | 0.33 (0.003) | 11.1 | 102,821 |
| | Testing | MDD age of onset | -0.24 (0.005) | 5.6 | 38,395 |

MSM-TRD: Maudsley Staging Method (MSM) treatment-resistant depression sum score. The p-value for testing in MSM-TRD is $1.18 \times 10^{-05}$ in the GLAD+.

**Table S9.** PPV, NPV, sensitivity and specificity for prediction models in external validation GLAD+ and UKB

| Cohort | Prediction score cut-off | 0.3 | 0.4 | 0.5 | 0.6 | 0.7 |
|---|---|---|---|---|---|---|
| **GLAD+ external validation** | PPV | 0.73 | 0.78 | **0.86** | 0.90 | 0.95 |
| | NPV | 0.82 | 0.72 | **0.58** | 0.51 | 0.41 |
| | Sensitivity | 0.98 | 0.93 | **0.77** | 0.64 | 0.38 |
| | Specificity | 0.17 | 0.42 | **0.72** | 0.84 | 0.96 |
| **UKB external validation** | PPV | 0.49 | 0.55 | **0.60** | 0.65 | 0.71 |
| | NPV | 0.81 | 0.78 | **0.76** | 0.73 | 0.71 |
| | Sensitivity | 0.79 | 0.66 | **0.56** | 0.46 | 0.33 |
| | Specificity | 0.53 | 0.69 | **0.79** | 0.86 | 0.92 |
| **UKB-resample external validation** | PPV | 0.77 | 0.81 | **0.85** | 0.87 | 0.90 |
| | NPV | 0.62 | 0.57 | **0.53** | 0.49 | 0.45 |
| | Sensitivity | 0.83 | 0.73 | **0.63** | 0.54 | 0.40 |
| | Specificity | 0.53 | 0.68 | **0.78** | 0.85 | 0.91 |

Note: GLAD+ results represent the model trained on UKB and tested for external validation in GLAD+. UKB and UKB-resample results represent the model trained on GLAD+ and tested for external validation in UKB and UKB resampled datasets.
UKB resample MDD prevalence 0.65
UKB MDD prevalence: 0.36
GLAD+ MDD prevalence: 0.69

**Table S10.** PPV, NPV, sensitivity and specificity for prediction models in GLAD+ and UKB

| Cohort | Prediction score cut-off | 0.3 | 0.4 | 0.5 | 0.6 | 0.7 |
|---|---|---|---|---|---|---|
| **GLAD+** | PPV | 0.81 | 0.85 | **0.89** | 0.91 | 0.93 |
| | NPV | 0.67 | 0.62 | **0.58** | 0.53 | 0.47 |
| | Sensitivity | 0.88 | 0.81 | **0.74** | 0.66 | 0.54 |
| | Specificity | 0.54 | 0.69 | **0.79** | 0.85 | 0.92 |
| **UKB** | PPV | 0.39 | 0.46 | **0.58** | 0.65 | 0.77 |
| | NPV | 0.87 | 0.83 | **0.77** | 0.73 | 0.68 |
| | Sensitivity | 0.96 | 0.85 | **0.60** | 0.45 | 0.22 |
| | Specificity | 0.15 | 0.43 | **0.75** | 0.86 | 0.96 |

PPV, positive predictive value, true positive / (true positive + false positive).
NPV, negative predictive value, true negative / (true negative + false negative).
Sensitivity, true positive / (true positive + false negative).
Specificity, true negative / (true negative + false positive).

**Table S11. Calibration for MDD prediction in GLAD+ and UKB**

| MDD prediction | | Brier score | Calibration slope | Calibration intercept |
|---|---|---|---|---|
| Train | Test | | | |
| GLAD+ | GLAD+ | 0.169 | 0.895 | 0.188 |
| UKB | UKB | 0.208 | 1.286 | -0.255 |
| GLAD+ | UKB | 0.197 | 0.793 | 0.038 |
| GLAD+ | UKB resample | 0.205 | 0.797 | 0.252 |
| UKB | GLAD+ | 0.174 | 1.399 | -0.114 |

According to the calibration slope and intercept, the model trained in GLAD+ and tested in the GLAD+, UKB, and UKB resample datasets is overconfident (slope < 1) and slightly underestimates risk (positive intercept). In contrast, the model trained in UKB and tested in the UKB and GLAD+ datasets is underconfident (slope > 1) and slightly overestimates risk (negative intercept). The Brier score ranges from 0.17 to 0.21. Since both GLAD+ and UKB have imbalanced samples, and major depression is a complex psychiatric disorder, the current Brier score is considered acceptable.

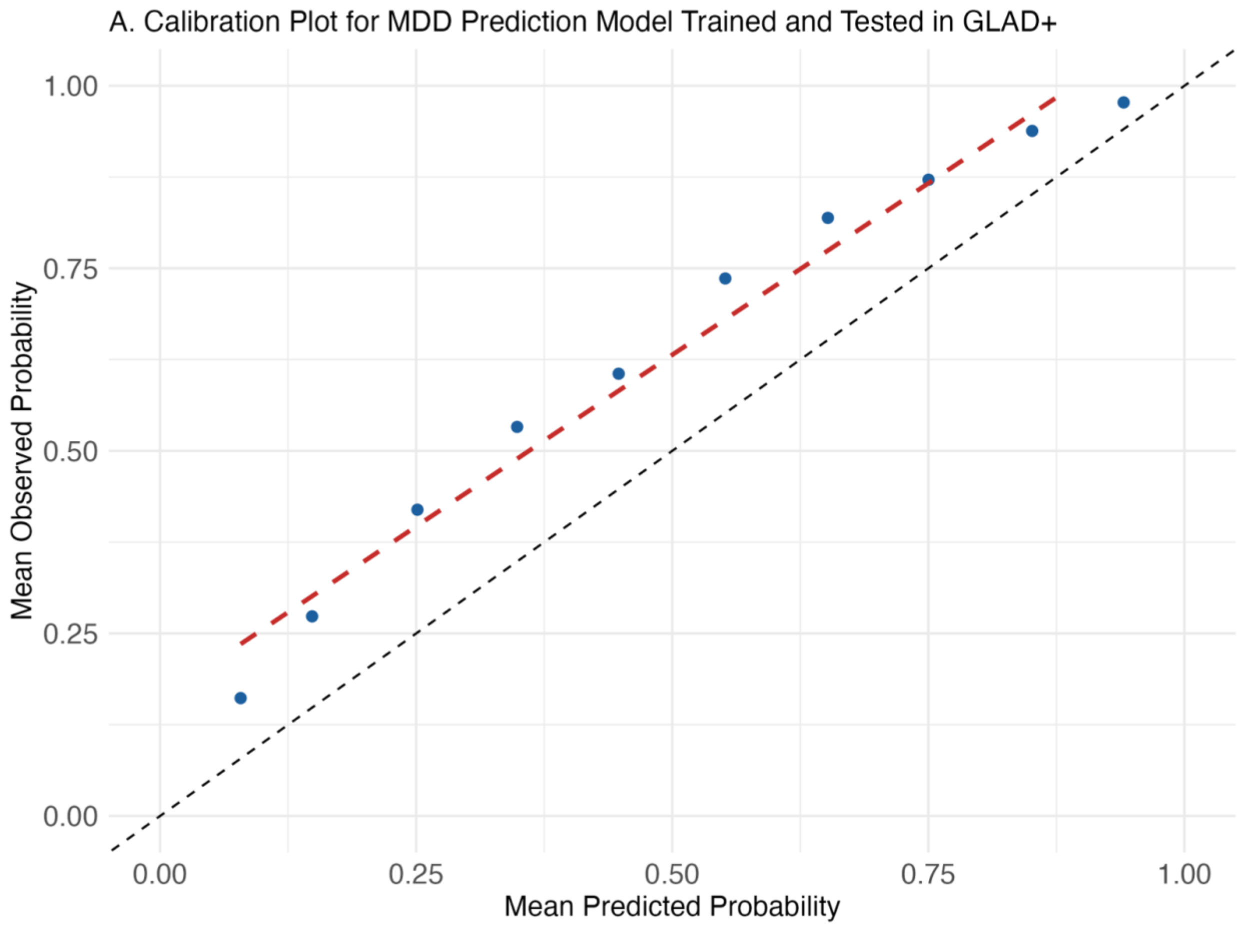
A. Calibration Plot for MDD Prediction Model Trained and Tested in GLAD+
Mean Observed Probability
1.00
0.75
0.50
0.25
0.00
0.00
0.25
0.50
0.75
1.00
Mean Predicted Probability

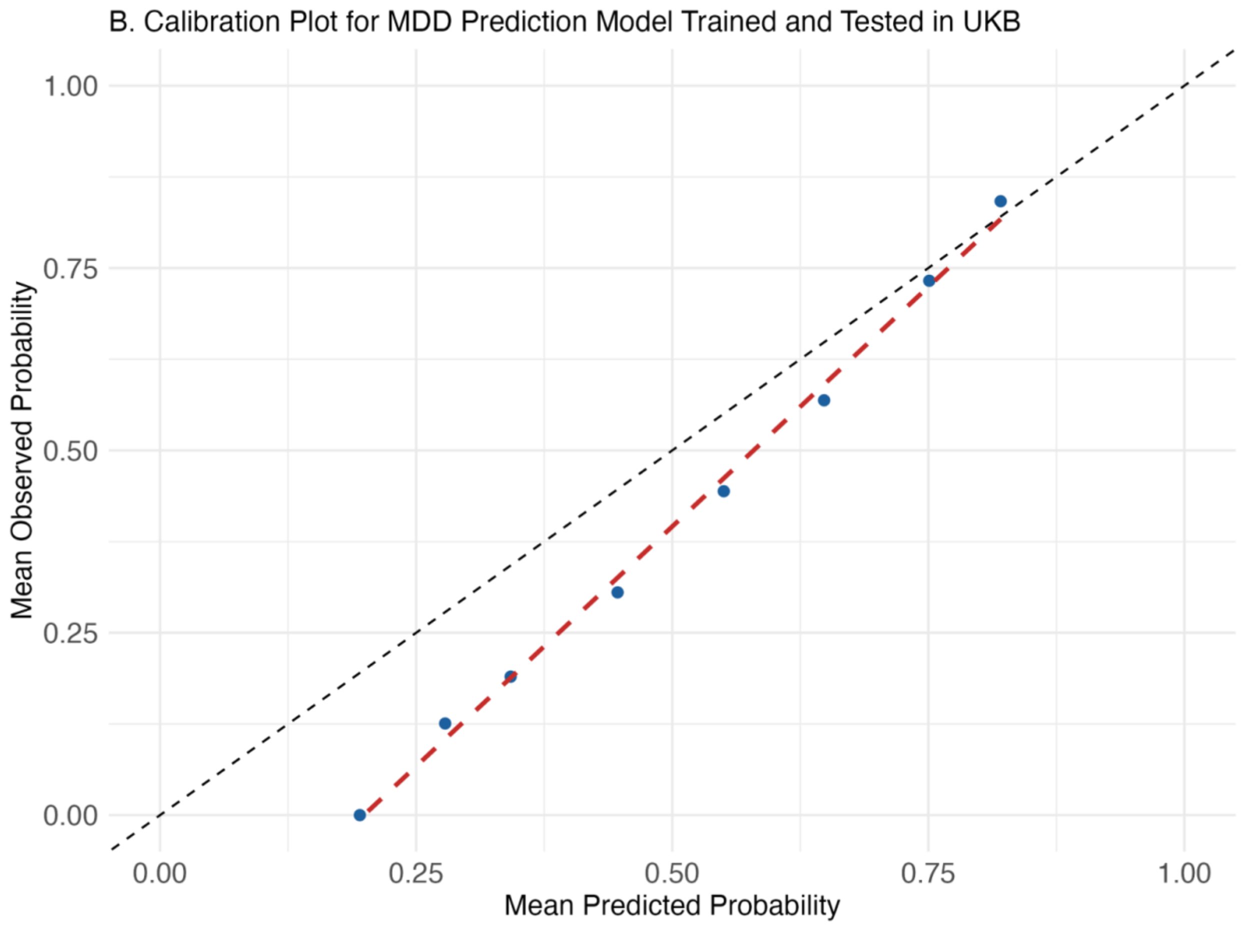
B. Calibration Plot for MDD Prediction Model Trained and Tested in UKB
Mean Observed Probability
1.00
0.75
0.50
0.25
0.00
0.00
0.25
0.50
0.75
1.00
Mean Predicted Probability

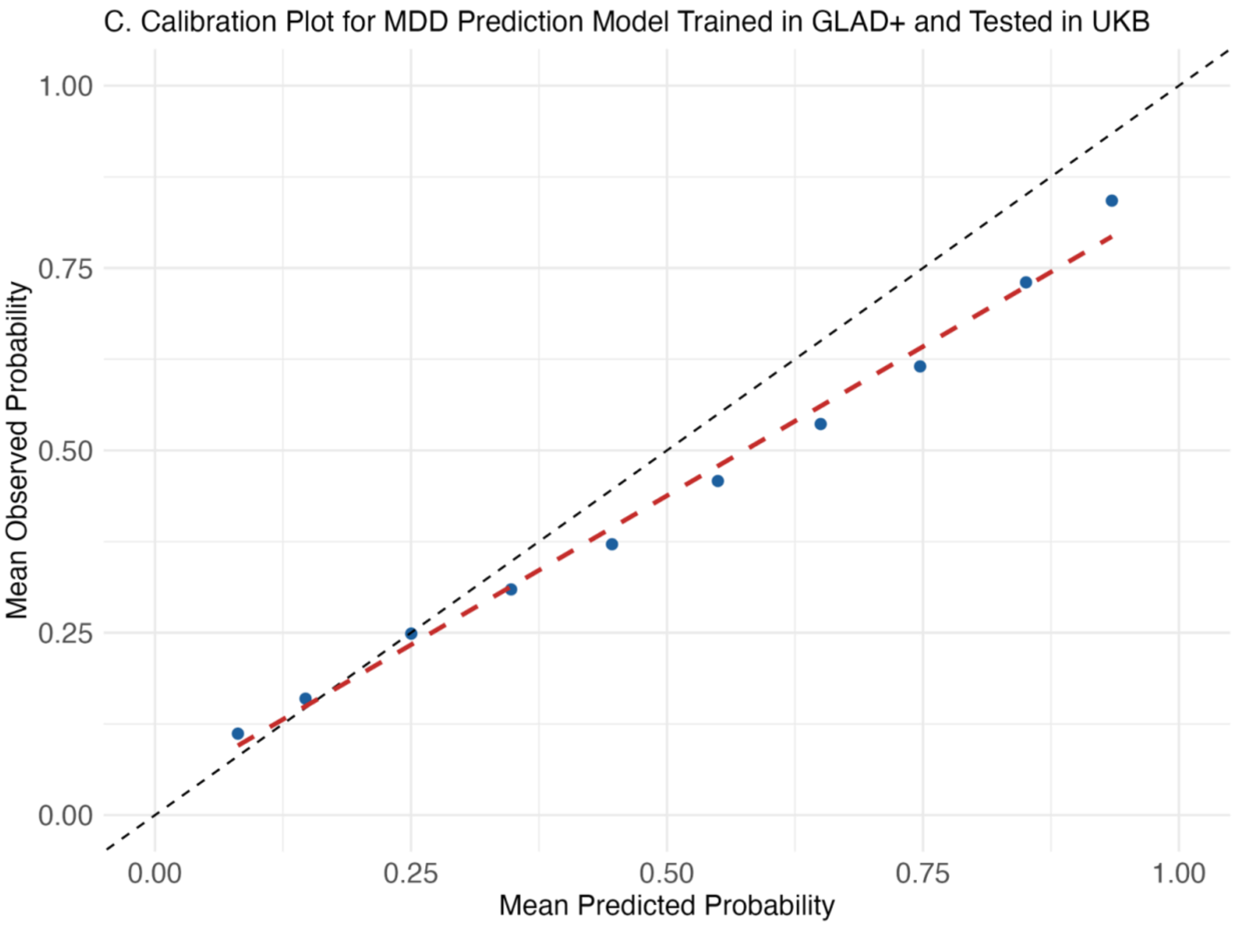
C. Calibration Plot for MDD Prediction Model Trained in GLAD+ and Tested in UKB
Mean Observed Probability
1.00
0.75
0.50
0.25
0.00
0.00
0.25
0.50
0.75
1.00
Mean Predicted Probability

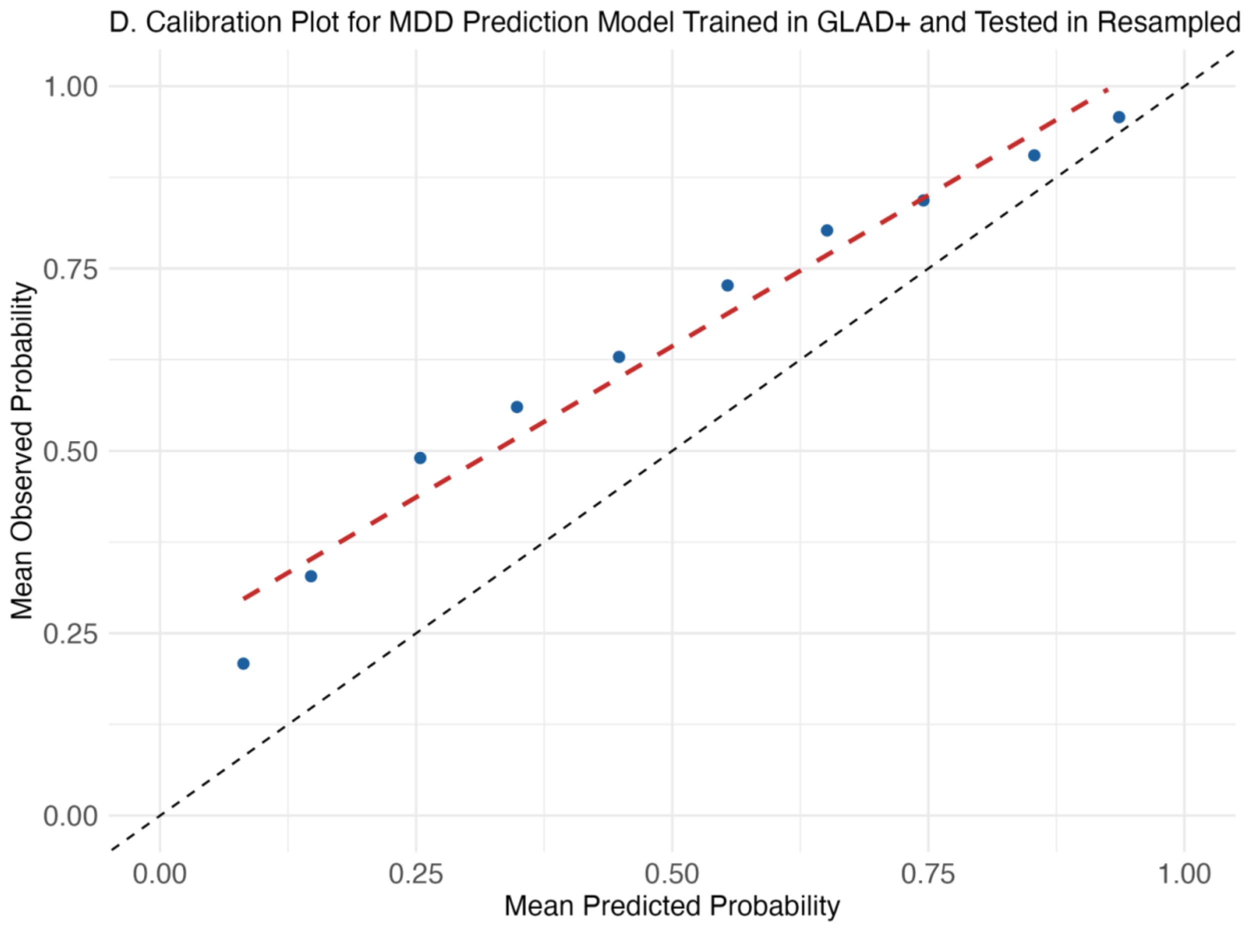
D. Calibration Plot for MDD Prediction Model Trained in GLAD+ and Tested in Resampled
Mean Observed Probability
1.00
0.75
0.50
0.25
0.00
0.00
0.25
0.50
0.75
1.00
Mean Predicted Probability

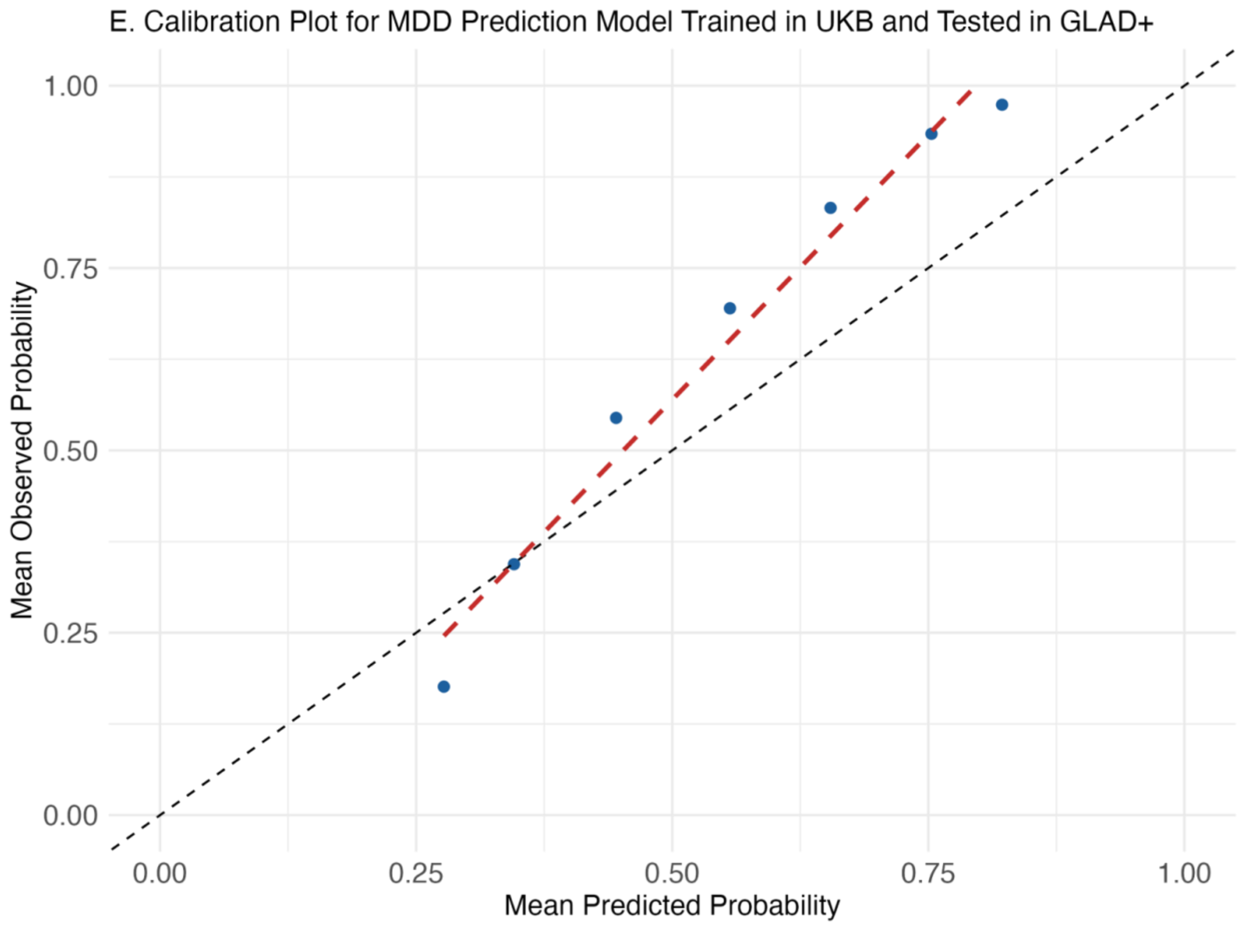


**Figure S7 A-E. Calibration plots for MDD prediction models in GLAD+ and UKB**

**Table S12.** The Pearson's correlations between top 5 predictors in GLAD+ and UKB

| Cohort | Predictor1 | Predictor2 | Pearson's correlation | 95% CI | P-value |
|---|---|---|---|---|---|
| **GLAD+** | Family history of depression | Childhood trauma | 0.17 | 0.15-0.19 | < 2.2e-16 |
| | Family history of depression | PRS_MDD | 0.15 | 0.13-0.16 | < 2.2e-16 |
| | Family history of depression | Family history of anxiety | 0.44 | 0.43-0.46 | < 2.2e-16 |
| | Family history of depression | Sex | 0.18 | 0.17-0.20 | < 2.2e-16 |
| | Childhood trauma | PRS_MDD | 0.16 | 0.14-0.18 | < 2.2e-16 |
| | Childhood trauma | Sex | 0.14 | 0.12-0.15 | < 2.2e-16 |
| | Childhood trauma | Family history of anxiety | 0.13 | 0.11-0.14 | < 2.2e-16 |
| | Family history of anxiety | PRS_MDD | 0.10 | 0.09-0.12 | < 2.2e-16 |
| | Family history of anxiety | Sex | 0.14 | 0.12-0.16 | < 2.2e-16 |
| | PRS_MDD | Sex | 0.08 | 0.06-0.09 | < 2.2e-16 |
| **UKB** | Family history of depression | Childhood trauma | 0.130 | 0.125-0.136 | < 2.2e-16 |
| | Family history of depression | PRS_MDD | 0.104 | 0.099-0.110 | < 2.2e-16 |
| | Family history of depression | Family history of anxiety | 0.266 | 0.261-0.272 | < 2.2e-16 |
| | Family history of depression | Sex | 0.148 | 0.142-0.153 | < 2.2e-16 |
| | Childhood trauma | PRS_MDD | 0.113 | 0.107-0.119 | < 2.2e-16 |
| | Childhood trauma | Sex | 0.073 | 0.068-0.079 | < 2.2e-16 |
| | Childhood trauma | Family history of anxiety | 0.074 | 0.069-0.080 | < 2.2e-16 |
| | Family history of anxiety | PRS_MDD | 0.067 | 0.061-0.073 | < 2.2e-16 |

| | | | | | |
|---|---|---|---|---|---|
| | Family history of anxiety | Sex | 0.095 | 0.089-0.101 | < 2.2e-16 |
| | PRS_MDD | Sex | 0.015 | 0.009-0.021 | 5.2e-07 |

The Pearson correlations among the top five predictors ranged from 0.08 (between PRS for MDD and sex) to 0.44 (between family history of depression and family history of anxiety) in GLAD+ and from 0.02 (between PRS for MDD and sex) to 0.27 (between family history of depression and family history of anxiety) in UKB (***Table S12***).

**Table S13**. leave out mPRS analysis

| Cohort | Dataset | Predictors | Liability $R^2$ | Liability $R^2$ deduction | AUC | Proportion of AUC deduction |
|---|---|---|---|---|---|---|
| **GLAD+** | Training | All predictors | 32.9% | | 0.84 | |
| | Training | Leave-out mPRS | 26.6% | 8.9% | 0.82 | 1.8% |
| | Testing | Leave-out mPRS | 29.1% | 0.34% | 0.81 | 2.4% |
| **UKB** | Training | All predictors | 23.1% | | 0.74 | |
| | Training | Leave-out mPRS | 20.8% | 7.6% | 0.73 | 1.4% |
| | Testing | Leave-out mPRS | 20.5% | 8.9% | 0.72 | 2.7% |

The testing dataset represents external dataset testing. For GLAD+ testing, the model was trained on the UKB and tested on the GLAD+ study, and vice versa.

#### The GLAD Study Group Authors

Gursharan Kalsi[1,2], Saakshi Kakar[1,2], Christopher Hübel[1,2,3,4], Ian Marsh[1,2], Laura H Meldrum[1,2], Iona Smith[1,2], Jahnavi Arora[1,2], Henry C. Rogers[1,2,5], Brett N. Adey[1,2], Zain Ahmad[1], Shannon Bristow[1,2], Charles J. Curtis[1,2], Susannah C. B. Curzons[1,2], Helena L. Davies[1,6,7], Molly R. Davies[8], Abigail R. ter Kuile[1,2], Sang Hyuck Lee[1,2], Yuhao Lin[1,2], Jared G. Maina[1,2], Monika McAtarsney-Kovacs[1,2], Dina Monssen[1,2], Jessica Mundy[1,2], Alish B. Palmos[1,2], Alicia J. Peel[1,2], Kirstin Purves[1,2], Christopher Rayner[1,2], Megan Skelton[1,2], Katherine N. Thompson[1,9], Rujia Wang[1,2], Johan Zvrskovec[1,2], Joshua E. J. Buckman[10], Ewan Carr[11], Antony J. Cleare[12,13], Katrina A. S. Davis[8], Kimberly A. Goldsmith[11], Colette R. Hirsch[14,15], Georgina Krebs[1,16], Donald M. Lyall[17], Katharine A. Rimes[12], Evangelos Vassos[1,2], David Veale[12,13], Janet Wingrove[18], Allan H. Young[19], Roland Zahn[19], Le Roy Dowey[20], Victor Gault[20], Chérie Armour[21], John R. Bradley[22], Ian R. Jones[23], Nathalie Kingston[22], Andrew M. McIntosh[24], Daniel J. Smith[25], James T. R. Walters[26], NIHR BioResource consortium, Jonathan R. I. Coleman[1,2], Matthew Hotopf[2,8], Thalia C. Eley[1,2], Gerome Breen[1,2]

*1. Social, Genetic, and Developmental Psychiatry Centre; Institute of Psychiatry, Psychology and Neuroscience; King's College London, London, UK*

*2. UK National Institute for Health Research (NIHR) Biomedical Research Centre, South London and Maudsley Hospital and King's College London, London, UK*

*3. National Centre for Register-based Research, Aarhus University, Aarhus, Denmark*

*4. Department of Pediatric Neurology, Charité - Universitätsmedizin Berlin, Berlin, Germany*

*5. Department of Psychiatry, Mount Sinai Health System, New York, USA*

*6. Mental Health Center Ballerup, Copenhagen University Hospital – Mental Health Services CPH, Center for Eating and feeding Disorders Research, Copenhagen, Denmark*

*7. Institute of Biological Psychiatry, Mental Health Center Sct. Hans, Mental Health Services Copenhagen, Roskilde, Denmark*

*8. Department of Psychological Medicine, Institute of Psychiatry, Psychology & Neuroscience, King's College London, London, UK*

*9. Department of Sociology, College of Liberal Arts, Purdue University, West Lafayette, IN, USA*

*10. CORE Data Lab, Centre for Outcomes Research and Effectiveness (CORE), Research Department of Clinical, Educational, and Health Psychology, UCL, London, UK*

*11. Department of Biostatistics and Health Informatics, Institute of Psychiatry, Psychology and Neuroscience, King's College London, London, UK*

*12. The Institute of Psychiatry, Psychology and Neuroscience, King's College London, London, UK*
*13. South London and Maudsley NHS Foundation Trust, Maudsley Hospital, London, UK*
*14. Department of Psychology, Institute of Psychiatry, Psychology and Neuroscience, King's College London, Denmark Hill, Camberwell, London, UK*
*15. Centre for Anxiety Disorders and Trauma, South London and Maudsley Hospital, London, UK*

*16. Research Department of Clinical, Educational and Health Psychology, University College London, London, UK*
*17. Institute of Health and Wellbeing, University of Glasgow, Glasgow, UK*

*18. Talking Therapies Southwark, South London and Maudsley NHS Foundation Trust, London, UK*

*19. South London and Maudsley NHS Foundation Trust and Centre for Affective Disorders, Department of Psychological Medicine, Institute of Psychiatry, Psychology, and Neuroscience, King's College London, London, UK*
*20. School of Biomedical Sciences, Ulster University, Coleraine, Northern Ireland, UK*

*21. Research Centre for Stress Trauma and Related Conditions (STARC), School of Psychology, Queen's University Belfast, Belfast, UK*

*22. University of Cambridge, Cambridge, UK*

*23. National Centre for Mental Health, Cardiff University, Cardiff, UK*

*24. Division of Psychiatry, Centre for Clinical Brain Sciences, University of Edinburgh, Edinburgh, UK*

*25. Division of Psychiatry, Centre for Clinical Brain Sciences, University of Edinburgh, Royal Edinburgh Hospital, Edinburgh, UK*
*26. Centre for Neuropsychiatric Genetics and Genomics, Division of Psychological Medicine and Clinical Neurosciences, School of Medicine, Cardiff University, Cardiff, UK*
